# The causal effects of chronic air pollution on the intensity of COVID-19 disease: Some answers are blowing in the wind

**DOI:** 10.1101/2021.04.28.21256146

**Authors:** Marc N. Conte, Matthew Gordon, Nicole A. Swartwood, Rachel Wilwerding, Chu A. (Alex) Yu

**Author notes:** Corresponding author: Department of Economics, Fordham University.

## Abstract

The threats posed by COVID-19 have catalyzed a search by researchers across multiple disciplines for policy-relevant findings about critical risk factors. We contribute to this effort by providing causal estimates of the link between increased chronic ambient pollutant concentrations and the intensity of COVID-19 disease, as measured by deaths and hospitalizations in New York City from March through August, 2020. Given concerns about unobservable characteristics that contribute to both ambient air pollutant concentrations and the impacts of COVID-19 disease, we instrument for pollutant concentrations using the time spent downwind of nearby highways and estimate key causal relationships using two-stage least squares models. The causal links between increases in concentrations of our traffic-related air pollutants (PM_2.5_, NO_2_, and NO) and COVID-19 deaths are much larger than the correlations presented in recent observational studies. We find that a 0.16 μg/m^3^ increase in average ambient PM_2.5_ concentration leads to an approximate 30% increase in COVID-19 deaths. This is the change in concentration associated with being downwind of a nearby highway. We see that this effect is mostly driven by residents with at least 75 years of age. In addition to emphasizing the importance of searching for causal relationships, our analysis highlights the value of increasing the density of pollution-monitoring networks and suggests potential benefits of further tightening of Clean Air Act amendments, as our estimated effects occur at concentrations well below thresholds set by the National Ambient Air Quality Standards.

---

Policymakers and researchers have sought to understand the drivers of differential impacts of COVID-19 disease across communities and demographic populations to mitigate its impacts. COVID-19’s impact on respiratory function and previous work on SARS and air-quality conditions (1), has led researchers to explore the relationship between air quality conditions and COVID-19 disease severity. One such study finds positive correlations between reduced chronic air quality and the severity of COVID-19 disease at the county level in the United States (2). Another suggests that air pollution is responsible for roughly 15% of the global mortality due to COVID-19 (3).

Direct mechanisms to explain how air pollution might influence COVID-19 disease severity include damage to the cilia (4) or overexpression of ACE-2 receptors (5). Potential indirect mechanisms include aggravation of pre-existing cardiovascular and respiratory conditions (4, 6). A survey of existing observational research suggests a correlation between COVID-19 mortality and both acute and chronic ambient concentrations of several compounds, including nitrogen dioxide and particulate matter (7). While acute exposures are sufficient to trigger these events, outcomes become more severe, often fatal, with chronic exposure.

However, these observational studies are unable to adequately address concerns that air quality and COVID-19 outcomes might be correlated with unobservable community and demographic characteristics. Our instrumental-variables approach for identifying the causal impact of chronic air-pollutant concentrations, here defined as a 10-year average preceding the pandemic, on COVID-19 outcomes among non-institutionalized individuals identifies substantially larger effects of increased ambient concentrations (e.g., PM_2.5_, NO_2_) on mortality than existing observational studies, even at concentrations well below threshold levels from the National Ambient Air Quality Standards.

## Challenges to Causal Identification

Work exploring links between chronic ambient pollutant concentrations and COVID-19 outcomes, including existing observational studies, is confronted by three challenges. First, aggregated ambient pollutant concentrations are typically relied upon as a proxy for exposure. This assumption is particularly strong given the coarse spatial resolution of most pollutant concentration data, whether derived from sparse governmental monitoring networks or satellite estimation.

Second, low-income and majority non-white neighborhoods are often subjected to higher concentrations of air pollutants (8, 9). Increased levels of particulate matter in these communities may be due to proximity to point sources of pollution (10, 11) or areas with traffic congestion (12). These existing findings suggest that poverty or other correlated characteristics could be confounding the results from recent observational studies about the relationship between air quality or race and outcomes of COVID infection (13).

Finally, the association between air quality and other demographic characteristics that impact health outcomes complicates efforts to identify the impact of chronic air-quality conditions on COVID-19 disease intensity in observational studies. Including demographic controls in these models does not solve this issue, as air pollution is causally linked with several demographic characteristics (e.g., income, pre-existing health conditions); the presence of these endogenous regressors will bias all coefficient estimates in such multivariate regressions (14). Importantly, it is difficult to know a priori the direction of the bias - correlational studies could either over or underestimate the effect of pollution on health.

We use quasi-experimental methods and detailed air-quality data from New York City, the one-time global epicenter of the pandemic (15), to causally identify the relationship between chronic air-quality conditions and the intensity of COVID-19 disease, as measured by mortality and hospitalizations between February 29 and August 30, 2020. We use two-stage least squares (2SLS) to estimate the causal link between ambient pollutant concentrations and COVID-19 outcomes, first finding the portion of air quality that results from being downwind of a highway, and then finding the effect of this exogenous variation in air quality on COVID-19 outcomes. Our estimation method compares COVID-19 outcomes in tracts within the same neighborhood that are within the same distance of a highway but that differ in the fraction of time spent downwind of the highway. We rely on the fact that the fraction of time downwind of a highway affects ambient air quality but is uncorrelated with any other individual or community characteristics that might affect COVID-19 outcomes (see figure 1 and supplement for details).

**Figure 1.**
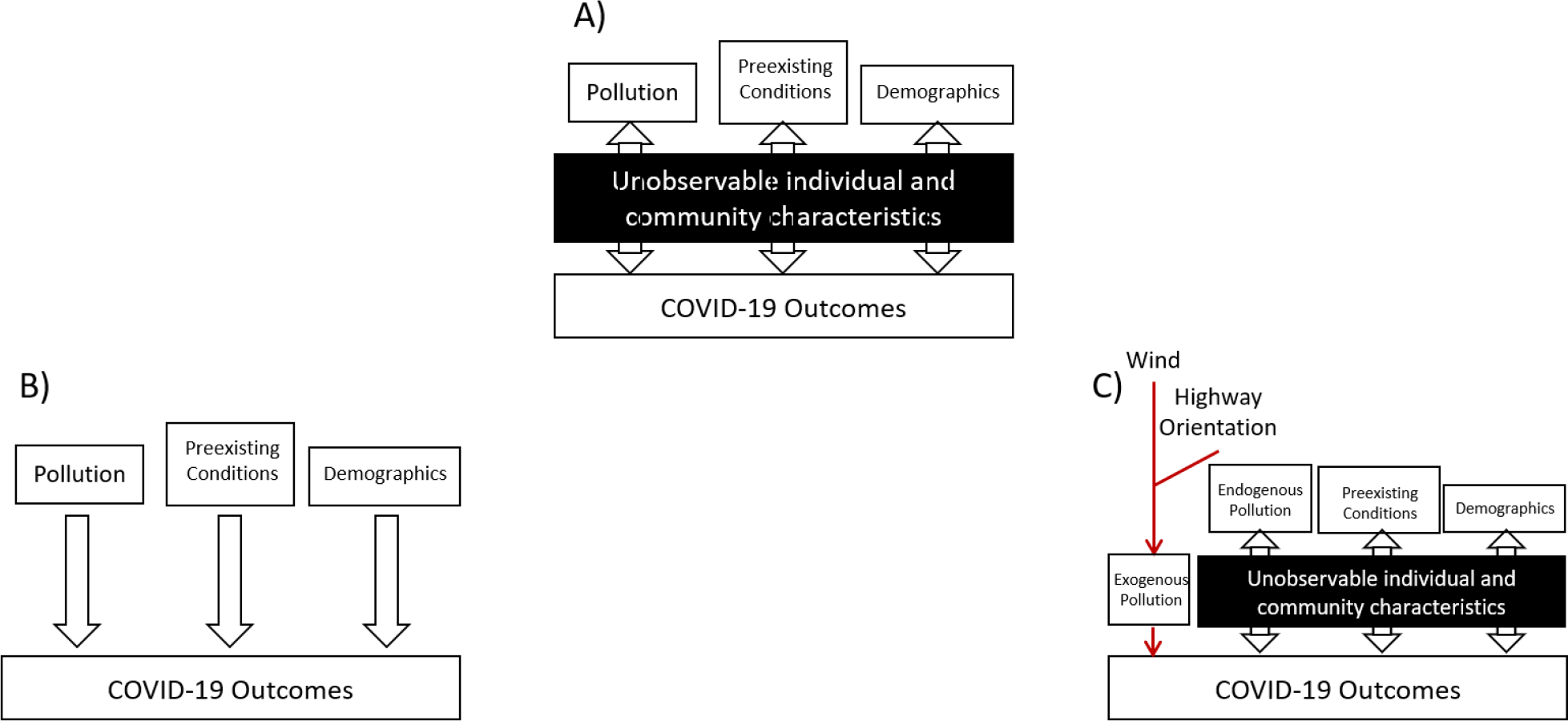
This figure displays the observed relationships between factors contributing to COVID-19 outcomes and how these relationships are estimated by alternative modeling approaches. A) presents the underlying, observed relationships, reflecting the endogeneous nature of several determinants of COVID-19 outcomes. B) illustrates how observational studies, which assume exogeneity of these variables, fail to account for their underlying relationships and provide biased coefficient estimates. C) illustrates our applications of an instrumental-variables approach, relying on the exogenous nature of wind characteristics and highway orientation, to identify the casual relationships between chronic ambient air quality and COVID-19 outcomes.

Figure 2 illustrates the challenge of designing an effective policy response to COVID-19 based on observational studies. While pollutant concentrations are highest in the wealthier, whiter parts of Manhattan, figure 2E illustrates that there was a significant decrease in the number of cell phone devices residing in these census tracts during the first wave of the pandemic, consistent with individuals in these neighborhoods leaving the city to avoid exposure to the disease. Figure 2F shows that the case rate, defined as the number of positive tests divided by the population of the census tract is substantially lower for this part of the city, potentially due to increased adoption of defensive behaviors.

**Figure 2.**
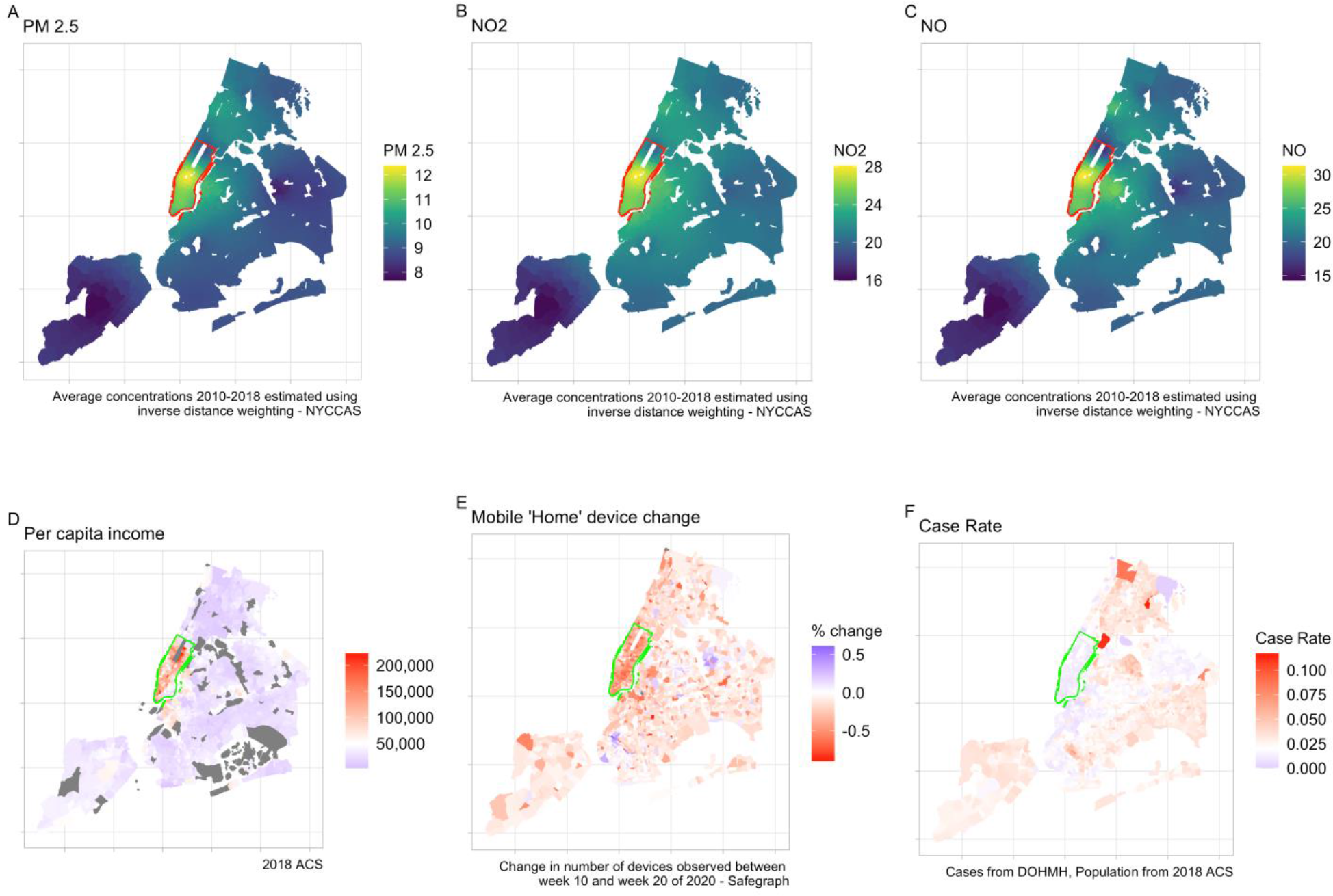
This figure depicts tract-level characteristics relating to chronic concentrations of PM_2.5_ (μg/m^3^) (a), NO_2_ (ppb) (b), NO (ppb) (c); 2018 per-capita income (d); the change in mobile devices based in each tract between week 10 and 20 of 2020 (e); and the case rate (positive test results divided by total tests within each tract) as of 08/31/2020 (f).

These data suggest that studies reliant on administrative data for population counts face measurement error in their rate-based dependent variables that is likely correlated with determinants of an individual’s susceptibility to the disease (e.g., income, ability to temporarily relocate), which would bias regression results with rate-based dependent variables. In fact, naive regressions of pollution on rate-based dependent variables show a negative relationship (see supplement Table S12). A rough attempt to account for these departures would effectively double death rates in this part of Manhattan, making them more comparable to those in the rest of the city (see supplement figure S3). For these reasons, we use log-transformed counts of hospitalizations and deaths, which, along with our 2SLS approach, can compare tracts with similarly sized populations without suffering from systematic measurement error in the dependent variable.

In the first stage of our estimation strategy, we identify the portion of pollutant concentrations that is solely due to the fraction of time spent downwind of highways, controlling for census tract distance to highways within neighborhoods. These regressions show that being continuously downwind of a nearby highway increases ambient concentrations of PM_2.5_ by 0.16 μg/m^3^, NO2 by 0.38 ppb and NO by 0.73 ppb relative to a tract in the same neighborhood that is equidistant from the highway but never downwind (see supplement table S2). These exogenous measures of ambient concentration are then used in our second-stage regressions to identify the causal impact of increased pollutant concentrations on our outcomes of COVID-19 disease.

## Results

Figure 3 presents the results from our instrumental-variable models showing the causal effects of long-term exposure to three pollutants on COVID-19 deaths (column 1) and hospitalizations (column 2). The figure panels illustrate results for three sets of observations: Citywide, Manhattan, which we restrict to tracts below 110^th^ Street, and Outer Boroughs, which includes tracts above 110th street in Manhattan. We also present results for several subsamples of tracts based on their distance to the nearest highway. For each set of observations, the figure presents the estimated coefficients and 95% confidence intervals for the focal pollutant on the logged count of our chosen measure of disease intensity.

**Figure 3.**
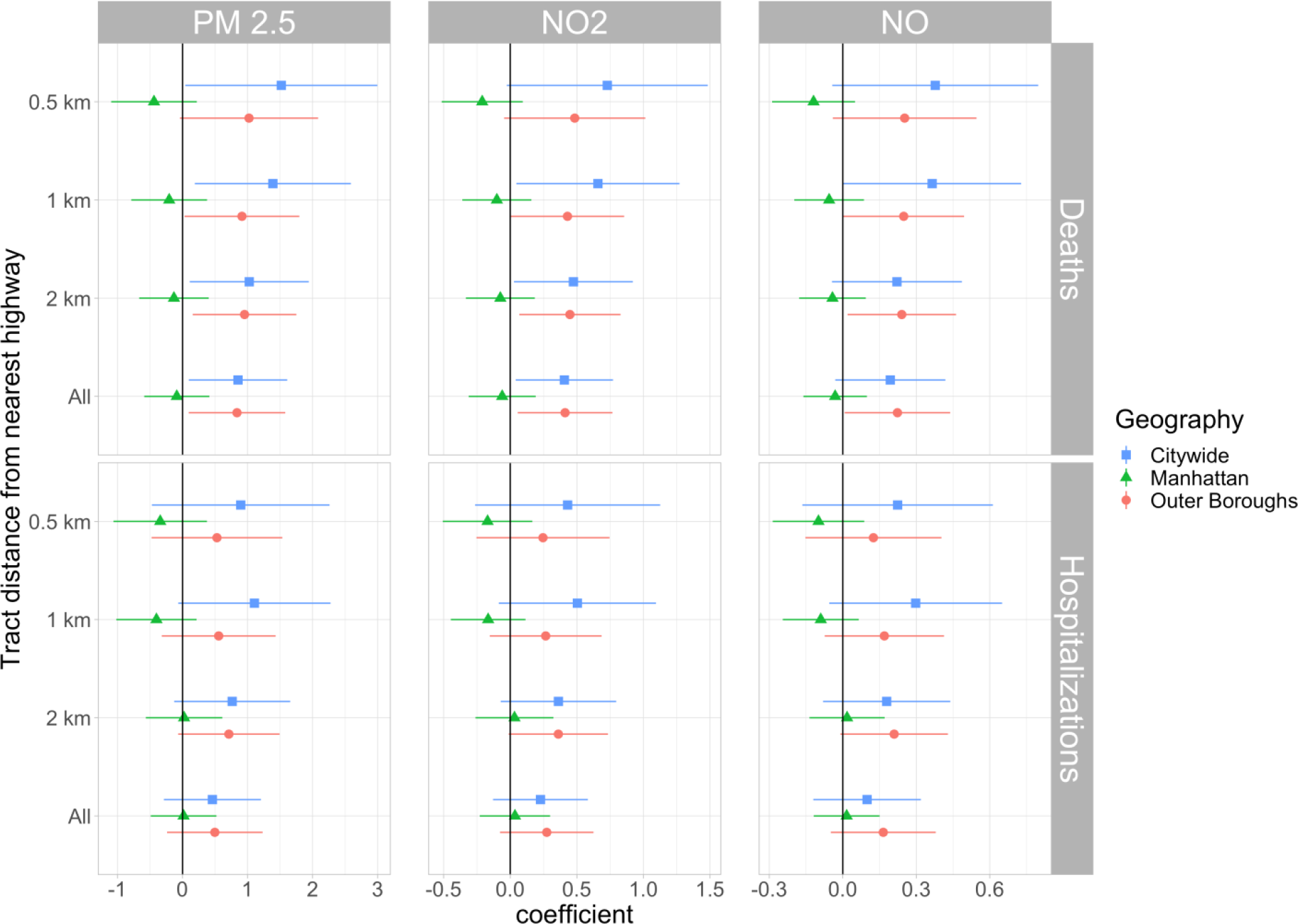
This figure presents the estimated coefficients and associated 95% confidence intervals on our instrumented measure of ambient pollutant concentration from the second-stage of our 2SLS log-linear models across our three geographic samples for tracts that lie within various distances of the nearest highway.

The causal impact of increased chronic pollutant concentration is positive for each of our considered pollutants, except in our Manhattan sample. The positive impact is consistently statistically significant in our models using both Citywide and Outer Boroughs samples for PM_2.5_ and NO_2_, although the confidence intervals for the subsample of tracts with centroids that lie within 0.5km of the nearest highway are larger than the rest, possibly due to a limited number of observations (only 842 out of our 2055 tracts lie within 0.5km of the nearest highway). The increasing coefficients in tracts that are closer to highways are consistent with results from our first-stage regressions showing that time downwind from highways within 1km of the centroid significantly predicts tract pollution, though the signal diminishes beyond that distance. The point estimates for NO are all positive, though the statistical significance is less consistent.

In our results for hospitalizations, we notice that the positive causal effect of increased TRAP concentration is never significant at the 5% level. However, the Outer Boroughs estimates are all significant at the 10% level. For the remainder of the paper, we report results from tracts that lie within 1km of a highway in our Outer Boroughs sample (results for other samples are presented in the supplement).

These results indicate that the increase in long term PM_2.5_ concentrations associated with being downwind of a nearby highway cause an average of 2.8 additional deaths (95% confidence interval [.4, 8.1]) per census tract (from a baseline average of 9.69 deaths per tract) and an increase of 5.33 hospitalizations [-.1, 16.0] (average 25.28 per tract). This is a 29% increase in deaths from a 0.16 μg/m^3^ increase in PM_2.5_ concentration – a much larger effect than existing research has shown.

Predicted deaths and predicted hospitalizations at various ambient pollutant concentrations based on our instrumental-variables models for all of our focal pollutants in our preferred sample, along with 95% confidence intervals, are presented in figure 4. The convexity of these curves is readily apparent, illustrating how quickly the average marginal effects of increased ambient pollutant concentrations on COVID-19 deaths increase across the observed range of concentrations, which lies well below National Ambient Air Quality Standards (NAAQS) for each of the three pollutants. Our non-linear model predicts that COVID-19 deaths increase sharply away from baseline levels as pollution increases. The average marginal effect of a 1 μg/m^3^ increase in PM_2.5_ (a 1.85 standard deviation increase in our sample) is to increase deaths by 160%.

**Figure 4.**
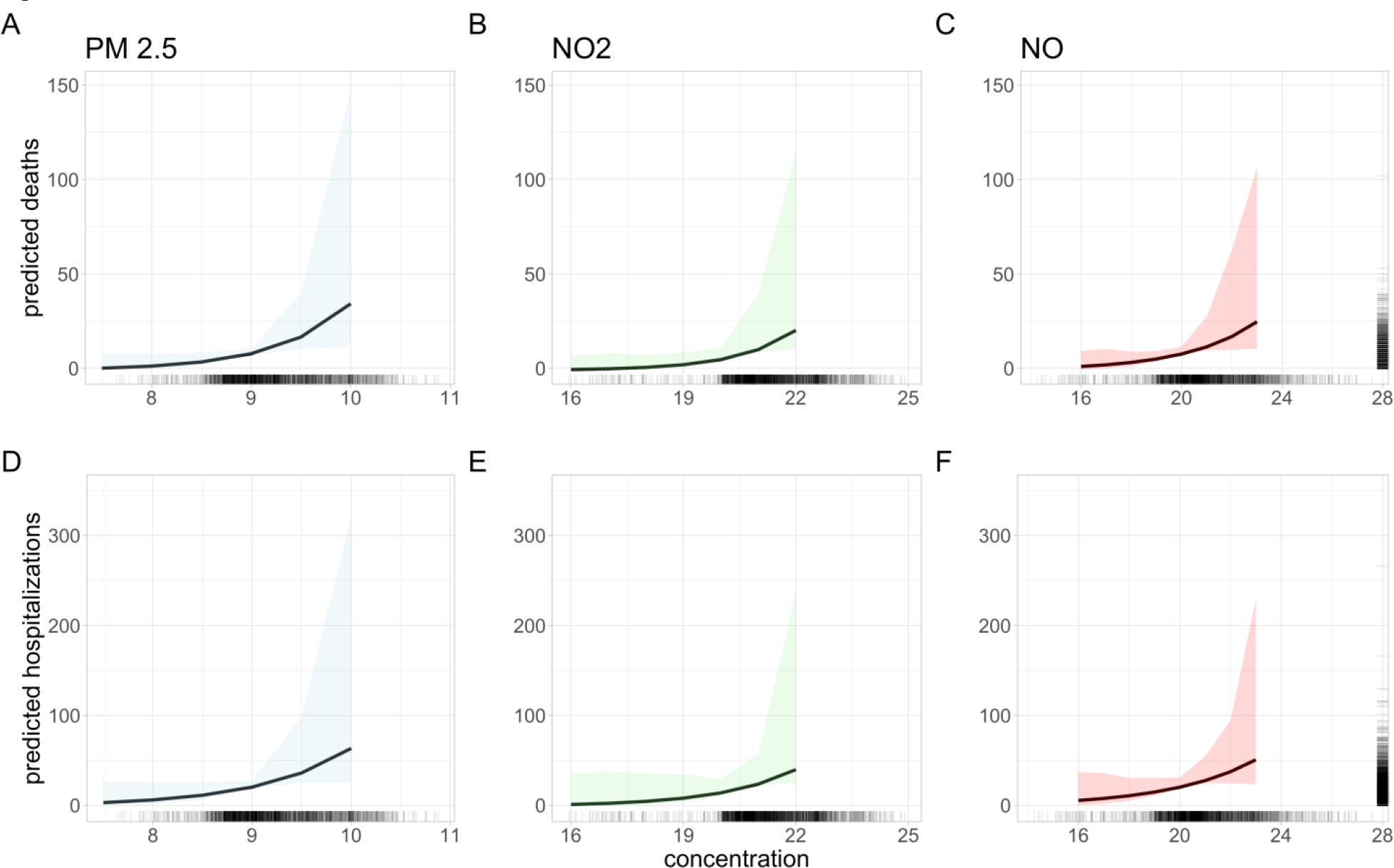
This figure depicts the predicted COVID-19 deaths (A) and hospitalizations (B) per census tract and associated 95% confidence interval based on the ambient concentration of our three focal pollutants: PM_2.5_ (1); NO_2_ (2); NO (3) in the set of tracts that lie within 1km of the nearest highway in our Outer Boroughs sample. The predicted outcomes are calculated from our 2SLS coefficient estimates at a subset of pollutant concentrations observed in the data (the density of concentrations is presented beneath the × axis in each plot). The confidence intervals are generated by taking the 2.5 and 97.5 percentiles of the empirical distributions of predicted deaths and predicted hospitalizations generated via 1,000 bootstrap iterations. The density of tract-level COVID-19 deaths and hospitalizations are presented on the y -axis at the far right of figures 4A and 4B, respectively.

Figures 5 and 6 present the causal effect of increased ambient pollutant concentration on the intensity of COVID-19 disease among specific racial and age groups, respectively. The plotted coefficients are estimated from models run on tracts that lie within 1km of the nearest highway, with at least 200 individuals of the given racial or age group. While the coefficients are similar across Asian/Pacific Islanders, Hispanic/Latino, and White individuals, only White individuals see a statistically significant increase in COVID-19 deaths with increasing ambient concentrations of our focal pollutants. The coefficient for Black/African American individuals appears to be a noisily-estimated zero, although the large standard errors on the estimated coefficient are notable. A similar pattern exists for the models of COVID-19 hospitalizations. The age results indicate that the effects of ambient air quality conditions on COVID-19 deaths are most pronounced for those individuals at least 75 years old, as might be expected. Though not statistically significant, it is a bit surprising that the point estimates for the 18-64 year-old age group are greater than those for the 65-74 year-old age group. Taken together, and knowing that the white population in New York City is on average older than the populations of the other racial groups considered, the higher death rates for whites in our Outer Borough sample may be due to the age distribution of this group of New York City residents.

**Figure 5.**
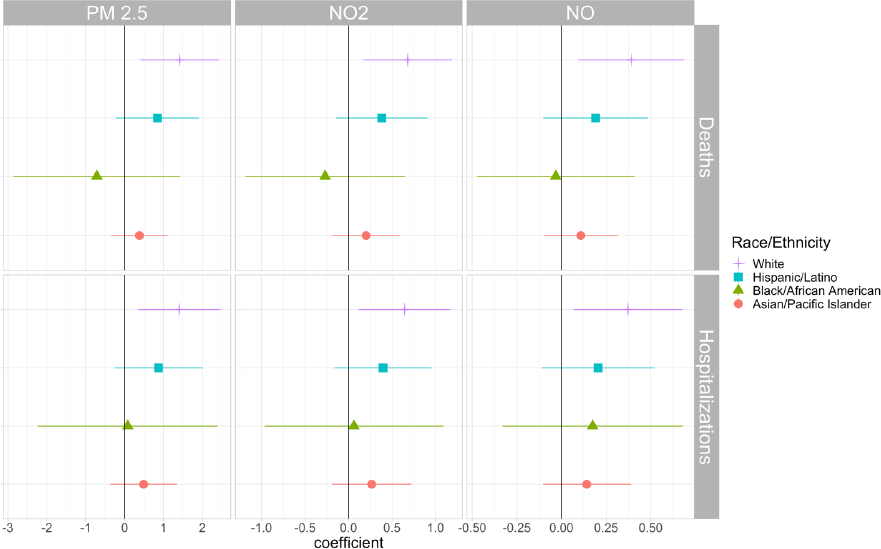
This figure presents the estimated coefficients and associated 95% confidence intervals from the second-stage of our 2SLS models that are stratified by Race/Ethnicity groups in the set of tracts that lie within 1km of the nearest highway in our Outer Boroughs sample on our instrumented measure of ambient pollutant concentration for our three pollutants: PM_2.5_; NO_2_; and NO.

**Figure 6.**
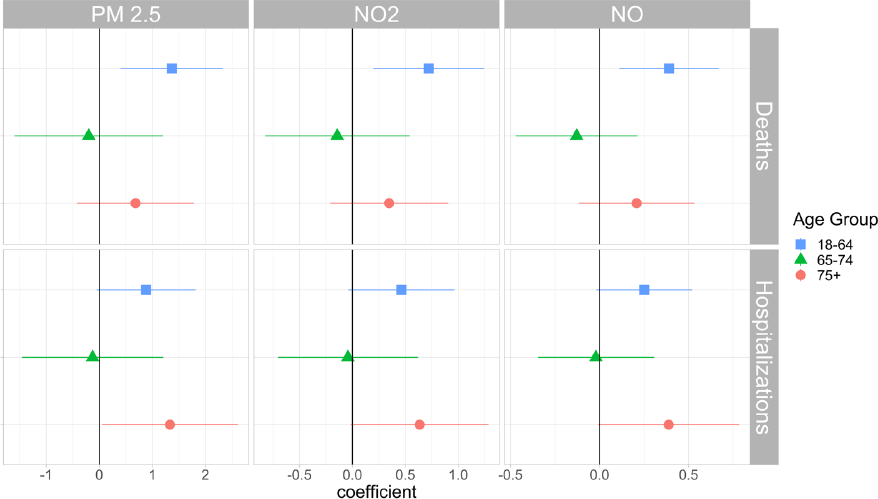
This figure presents the estimated coefficients and associated 95% confidence intervals from the second-stage of our 2SLS models that are stratified by Age groups in the set of tracts that lie within 1km of the nearest highway in our Outer Boroughs sample on our instrumented measure of ambient pollutant concentration for our three pollutants: PM_2.5_; NO_2_; and NO.

## Discussion

We have identified the causal relationship between chronic ambient concentrations of several air pollutants and the intensity of COVID-19 disease. Using an instrumental-variable approach to develop measures of chronic ambient air quality that are uncorrelated with characteristics that might confound causal inference, we find that increases in the average chronic concentration of three traffic-related air pollutants lead to statistically significant and surprisingly large increases in deaths due to COVID-19. The results of our main models predict a 29% increase in COVID deaths for a 0.16 μg/m^3^ increase in the ambient concentration of PM_2.5_.

The estimated magnitudes in our causal model of COVID-19 deaths are much larger than those reported in recent observational studies looking at chronic ambient pollutant concentrations, emphasizing the importance of our instrumental-variables approach. Support for such large impacts of air quality conditions on COVID-19 outcomes comes from statistics compiled by the NYC DOHMH showing that individuals with underlying conditions, including asthma, cancer, diabetes, and hypertension, have a COVID-19 case fatality rate (CFR) roughly 100x that of individuals without such underlying conditions (16). Given the association between air pollution and the development of chronic health conditions as well as sub-clinical deterioration of cardio-respiratory function (17, 18), a moderate increase in the prevalence of these underlying conditions could explain the apparently large effect on deaths. These correlations also suggest why studies that control for underlying health conditions might underestimate the true effect of air pollution on COVID-19 outcomes: if chronic air pollution led to the development of underlying conditions, controlling for these conditions would bias the estimated effect of air pollution toward zero.

Our estimated effects are observed at chronic ambient pollutant concentrations that fall below the NAAQS thresholds for PM_2.5_ (three-year annual average of 12 μg/m^3^ for the primary standard) and NO_2_ (annual average of 53 ppb). Our findings suggest that further amendment of the Clean Air Act could increase its net benefits to society, which are already known to be substantial (19).

There are a number of possible explanations for the lower statistical significance of the response of COVID-19 hospitalizations to chronic air-quality conditions, including the possibility that the well-publicized bed shortage in hospitals kept potential patients with mild symptoms from seeking care. Another potential explanation is that our variable, defined as a hospitalization within 14 days of the diagnosis date (before or after) is simply an imprecise measure of COVID-19 hospitalizations, reducing the statistical significance, though not the magnitude of the relationship.

Our roughly equal coefficients across racial and ethnic groups indicate that studies that find significant differences in outcomes between groups are likely failing to account for unobservable determinants of pollutant exposure. Our imprecisely estimated effects for Black/African American residents seem to be consistent with the different age distributions across the races as well as the fact that the distribution of Black residents of New York City is much less uniform across census tracts than that of residents of other races. The unexpected pattern of impacts across age groups might be explained by the simple fact that there are more New Yorkers in the 18-64 year-old age group than the older, presumably more susceptible, 65-74 year-old group. One alternative explanation is survivorship bias. In order to survive to age 65-74, one must be healthy enough to survive other mortality risks, such as heart disease, diabetes, etc. Due to a lack of these underlying conditions, while these persons might be at increased risk of infection, they might experience less severe COVID-19 disease due to their overall health status. Over 75 years of age, immunosenescence might be so severe that general health status is less important.

Our analysis highlights several additional issues relating to the unequal impacts of COVID-19 with relevance to researchers and policymakers alike. First, different populations have different access to defensive actions that can limit exposure to COVID-19. Failure to account for differences in the ability to temporarily move away from infection hotspots can lead to systematic measurement error in commonly-used, rate-based measures of COVID-19 outcomes that can bias regression results from both observational and causal studies. The findings from our age and racial group analyses seem to confirm the importance of defensive behaviors and relative exposure as explanations for differential impacts of COVID-19 disease by race rather than physiological differences. While the spatial patterns of air quality and demographic characteristics in New York City differ from other parts of the nation, our findings reinforce the point that disparities exist across incomes and racial groups regarding the ability to mitigate exposure to environmental hazards. Our findings suggest that the wealthier, whiter population of Manhattan below 110^th^ Street likely avoided significant increases in COVID deaths and hospitalizations by adopting defensive behaviors that were unavailable to other residents.

We emphasize that our estimate of chronic air-quality conditions does not fully capture the cumulative impact of ten years of exposure to our three air pollutants of interest. Rather, we have identified the causal relationship between average ambient air-quality conditions and COVID-19 outcomes. This limitation relates to the challenge of proxying for exposure to pollutants with measures of their ambient concentration, as the ability to moderate exposure may differ across individuals. This is a challenge for all existing work that has explored the health effects of air quality and their implications for behavior.

Access to pollutant concentrations at a fine spatial scale through the NYCCAS monitoring network was essential for our project to avoid the limitations of the EPA monitoring network and concentration estimates derived from satellite measurements (20-23). New York City is one of the few places in the United States with such a concentrated network of monitors, and our results suggest that there is great value in the establishment of a national network at similar scales.

Still, the large confidence intervals around our point estimates may be partially explained by the presence of numerous local pollutant sources in New York City, the rapid decay of TRAP with increasing distance from highways, and an insufficiently dense monitoring network, further emphasizing the importance of improved data in generating precisely-estimated relationships. The availability of localized information would increase the salience of pollutant levels, possibly allowing for increased uptake of defensive behaviors regarding pollutant exposure. We have shown these behaviors to be very important in the context of COVID-19, which is just one of the respiratory ailments through which air-pollutant exposure might lead to premature death and reduced quality of life.

## Data and Methods

Conducting our study in a relatively small spatial area allows us to avoid issues that challenge cross-sectional observational studies conducted over larger regions (e.g., national studies), such as correlation between air pollutant concentrations and employment opportunities, as well as to take advantage of a dense local pollution monitoring network.

We use the New York City Community Air Survey (NYCCAS) monitor readings for our concentration levels of the three considered pollutants and calculate the 10-year average concentration of each pollutant from 2009-2018, which is slightly shorter than the average of the median years that individuals have lived in their current residents across the city (11.7). There were between 66 and 110 NYCCAS monitors active between 2009 and 2018, as compared to the 12 EPA monitors in the city, allowing us to avoid several issues associated with the sparse and strategically-sited EPA monitoring network (20,23) and satellite-derived air-quality data that are downward biased (21), calibrated to ground-based networks (22), and too coarse to detect our observed relationships given the rapid decay of ambient concentrations of TRAP with increasing highway distance. Wind data (direction, speed) comes from hourly observations for all days 2009-2018 from the National Centers for Environmental Information Integrated Surface Database from the 4 closest weather stations at NYC-Area Airports (JFK, LaGuardia, Newark, and Teterboro). Our census-tract level data on COVID-19 mortality and hospitalizations comes from the New York City Department of Health and Mental Hygiene. We use cell phone data from Safegraph to develop measures of flows into and out of census tracts during the peak of the epidemic in New York City. Finally, demographic information at the tract level is taken from the 5-year 2018 American Community Survey.

The recent economic literature on the health effects of acute air-quality conditions includes efforts to accommodate the endogeneity of air-quality using weather-related variables. Weather-related variables provide identifying variation in pollution that is unlikely to be correlated with unobserved determinants of health outcomes (24-26). We build on this existing approach in two ways. First, we use NYCCAS data to create inverse-distance-weighted measures of chronic air-quality conditions for each census tract. Second, we use ten years of hourly wind direction data to create measures of the fraction of time that each census tract spends downwind of highways, acknowledging that the effects of being downwind vary with distance to the nearest highway.

We use a two-stage least-squares (2SLS) econometric framework to identify the causal impact of chronic air quality conditions on the intensity of COVID-19 infection in New York City, using the fraction of time spent downwind of highways at different distances from the census tract (e.g., 0.5km, 1km, etc.) and wind speed as instruments for chronic air quality conditions, based on average concentration of each pollutant over the 10-year period from 2009-2018. Our identification strategy compares tracts within the same neighborhood that lie within the same distance of the highway, but in different directions.

Our second-stage regressions estimate the causal impact of a change in pollutant concentration on two different measures of the intensity of COVID-19 infection: hospitalizations and deaths. We run regressions on the log-transformed count data. We also run Poisson regressions on dependent count variables and obtain similar estimates of the marginal effects of each of our pollutants (see supplement for these details).

## Data Availability

Per a data-use agreement with the NYC DOHMH, we cannot share data on COVID-19 outcomes or testing.

