## Supplementary Materials for "The causal effects of chronic air pollution on the intensity of COVID-19 disease: Some answers are blowing in the wind"

April 27, 2021

[†]Department of Environmental Studies, Yale University
[‡]T.H. Chan School of Public Health, Harvard University
[§]Department of Economics, Fordham University
[¶]Department of Economics, Wake Forest University

**4 Supplement References** **46**

### 1 Data

Our analysis is based on data from a number of different sources. The census-tract level data on COVID-19 cases, hospitalizations, and death counts comes from the New York City Department of Health and Mental Hygiene, covering all census tracts in New York City (NYC) from February 29, 2020 to August 30, 2020. The data set includes 18,856 deaths and 48,427 hospitalizations stratified by census tract, race, and age.

We use data from the New York City Community Air Survey (NYCCAS) to estimate ambient concentrations of traffic related air pollution (TRAP), focusing on $PM_{2.5}$, $NO_2$, and NO, from 2009-2018. NYCCAS measured ambient pollutant concentrations from 66-110 monitors placed around NYC over this time period. NYCCAS monitors record pollutant concentrations at a much finer spatial scale than the EPA. Figure S1 plots the $PM_{2.5}$ monitoring network for both NYCCAS and EPA monitors. We take bi-weekly pollution concentrations measured at each monitoring site and calculate 10-year average pollutant concentration for each census tract centroid in NYC, using squared inverse distance weighted (IDW) averages. Our results are also robust to proxying for exposure with average ambient pollution measured from the closest monitor; however, this noisier measure results in wider confidence intervals for our estimates, especially in Manhattan (see Table S14, panel C).

We collect hourly wind speed and wind direction data from the National Centers for Environmental Information (NCEI) Integrated Surface Database (ISD) from the 4 closest weather stations at NYC-area airports. We use this data to develop a count of the hours during our study period that a census tract is downwind of the nearest highway to calculate the fraction of time a census tract is downwind of a highway over a 10-year period. Due to anomalies in direction and speed, in addition to periods of missing data, we omit data from the station in Central Park and rely exclusively on readings from NYC-area airports. When constructing our measures from the closest monitor to each census tract, we rely on data from JFK, Newark, La Guardia, and Teterboro airports. Histograms of hourly wind directions are shown in Figure S2.

We incorporate cell phone mobility data from Safegraph to examine avoidance behavior. Safegraph collects location data from 45 million mobile devices and provides aggregated statistics at the census block group level on the amount of time that mobile

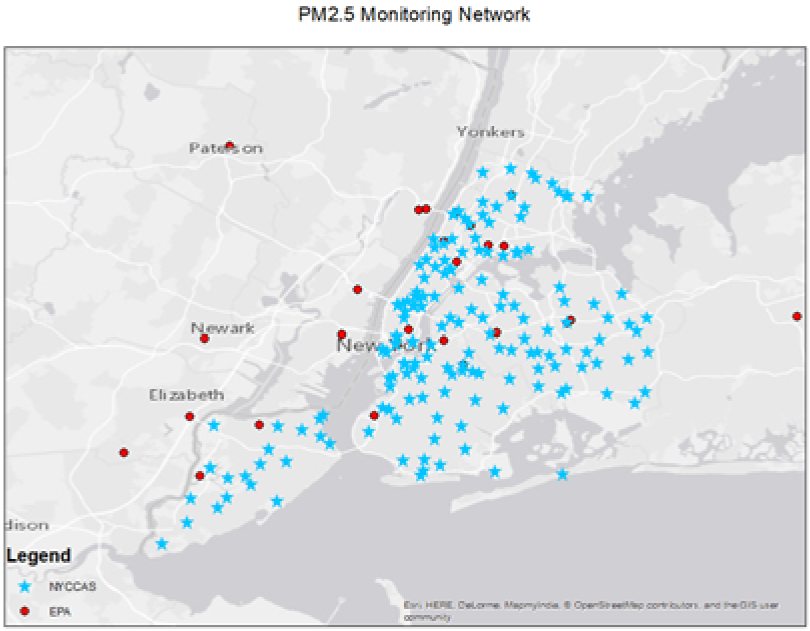

Figure S1: $PM_{2.5}$ Monitoring Network in NYC

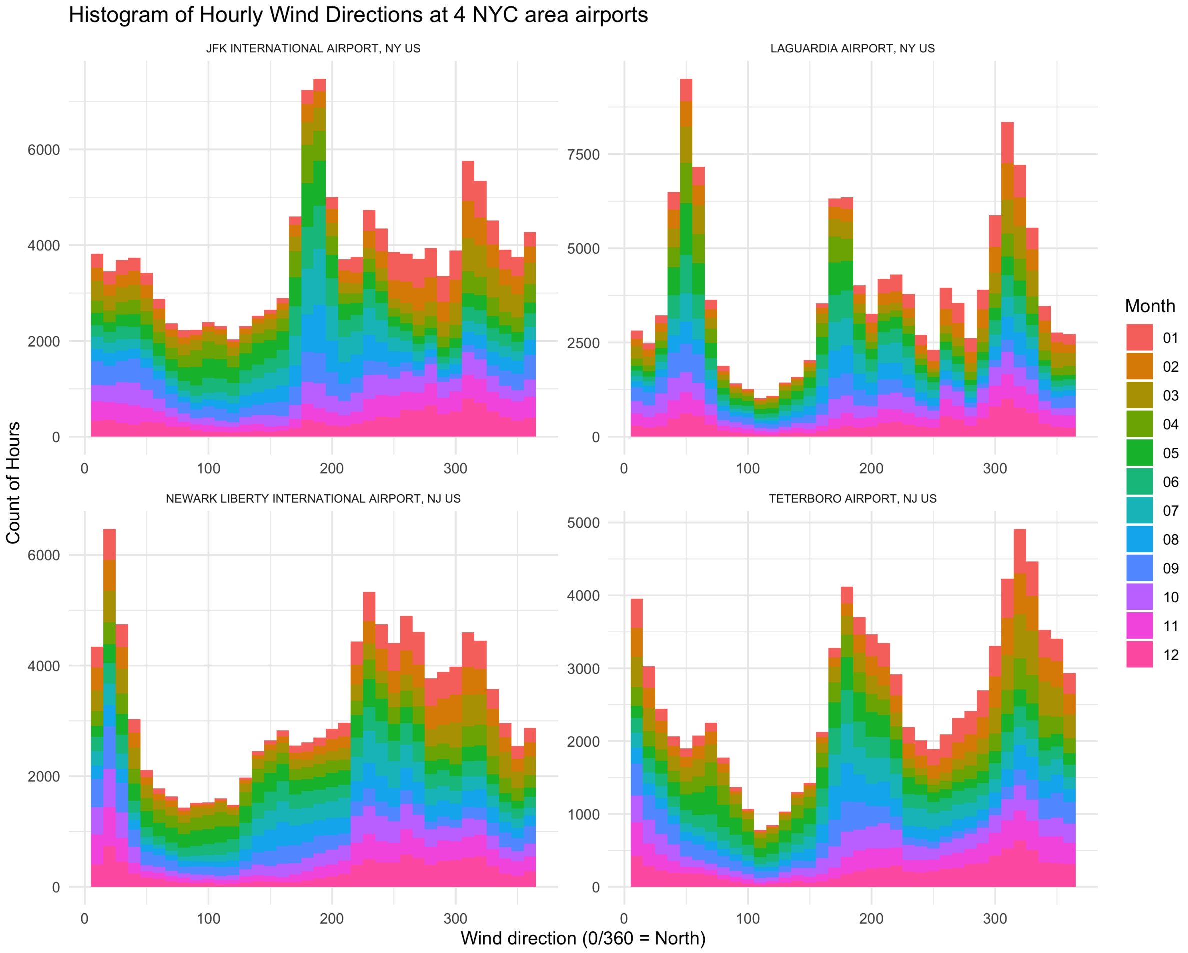

Figure S2: Histogram of Hourly Wind Directions at 4 NYC-area Airports devices are in the home, outside of the home, and engaged in work behavior, as well as distance traveled. The data are aggregated to the census tract level to match with our outcome variables. Figure S3 shows the importance of this behavior, calculating COVID-19 death and hospitalization rates using 2018 ACS population in the denominator (panels A and D) compared to using population adjusted for the number of devices that left the city between March 8 and May 11, 2020 (week 10 and 20 of the year), which corresponds to the peak of the initial wave of COVID-19 in the city (panels B and E). Blue points in the scatterplots (panels C and F) show the importance of measurement error in census tract population - addressing this measurement error with cellphone mobility data results in much higher death and hospitalization rates in Manhattan relative to the rest of the city (see Table S12, panels A and B). This is further discussed in Section 3.

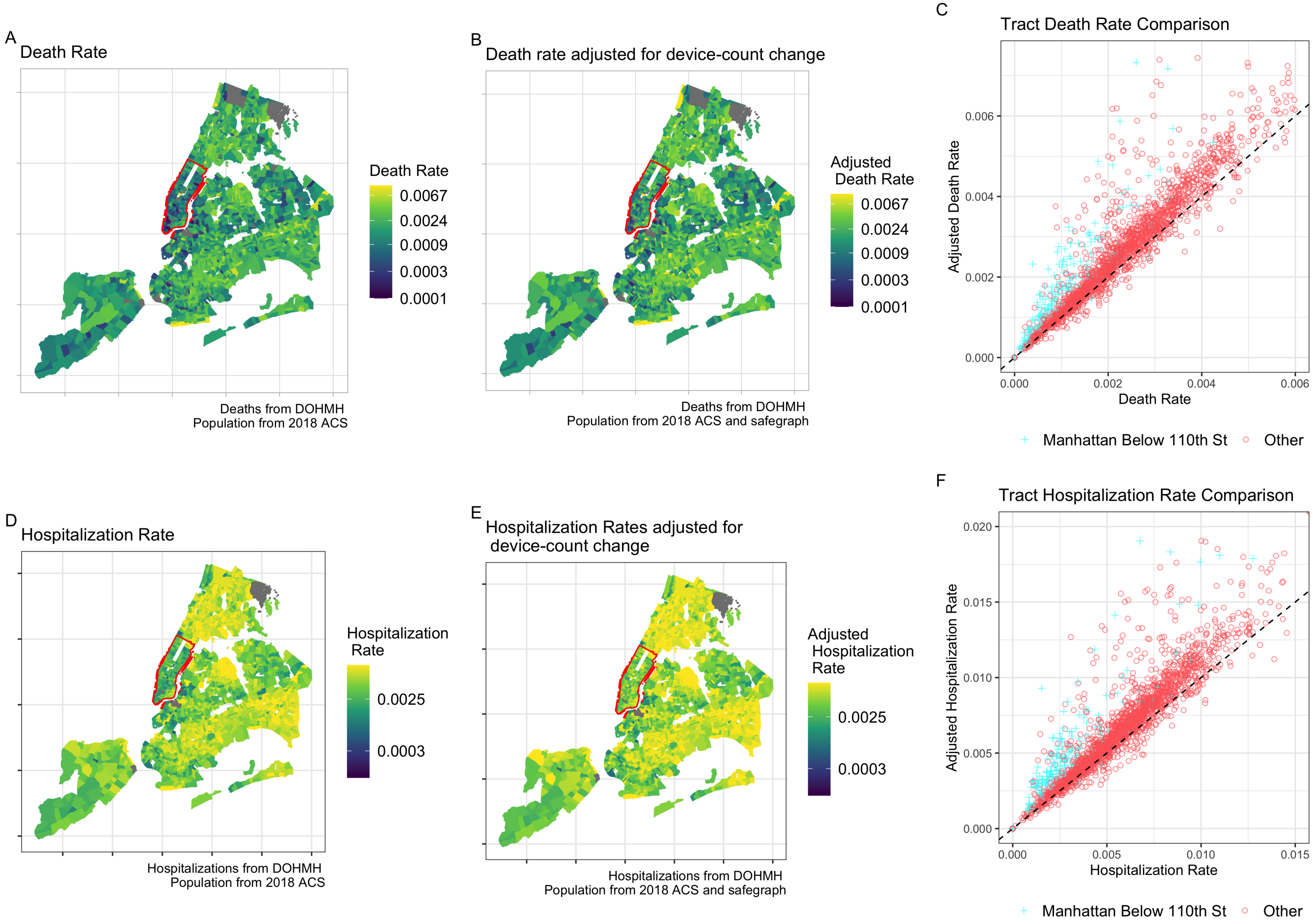

Figure S3: Death and Hospitalization Rates adjusted for departures

Our socioeconomic and demographic variables come from the 2018 5-year American Community Survey (ACS) and are used in some of our robustness checks. In our main specifications, we use tracts with more than 500 people. Our results are robust to using a threshold population of 100 as well (see Table S14, panel A). Average tract population is 3,900 individuals. 68 out of 2,165 tracts have fewer than 500 people and 55 have fewer than 100 people. Most of these tracts are parks or non-residential areas.

To generate exogenous measures of our focal TRAP components, we use information about each tract's orientation to the highway network in NYC. GIS data on NYC's highways comes from TIGER/Line Shapefiles (2015) for primary and secondary roads in NYC. Our interest is in a census tract's position relative to highway segments in NYC. To obtain accurate measurements of the distance and direction from each census tract centroid to surrounding highways, we divide the city into a grid with .01 by .01 degree cells (approximately 1.11 km on each side). Any lengths of highways within each grid cell are considered a 'highway segment'.

Summary statistics are presented for all tracts and for our main sample (Outer Borough - between .05km and 1km from a highway) in Table S1.

#### Table S1: Summary Statistics

##### A. Citywide - > 0.05 km from Nearest Highway with Population > 500

| Statistic | N | Mean | St. Dev. | Min | Pctl(25) | Pctl(75) | Max |
|---|---|---|---|---|---|---|---|
| Tract Population | 2,055 | 4,033.02 | 2,178.56 | 514 | 2,470 | 5,074 | 28,272 |
| Downwind 3-5km (%) | 2,055 | 0.75 | 0.16 | 0.00 | 0.66 | 0.88 | 0.92 |
| Downwind 2-3km (%) | 2,055 | 0.57 | 0.21 | 0.00 | 0.41 | 0.74 | 0.92 |
| Downwind 1-2km (%) | 2,055 | 0.43 | 0.23 | 0 | 0.3 | 0.6 | 1 |
| Downwind 0.5-1km (%) | 2,055 | 0.18 | 0.18 | 0 | 0 | 0.3 | 1 |
| Downwind $<$ 0.5km (%) | 2,055 | 0.10 | 0.14 | 0 | 0 | 0.2 | 1 |
| Distance to Closest Highway | 2,055 | 0.84 | 0.70 | 0.05 | 0.30 | 1.18 | 5.19 |
| $PM_{2.5}$ | 2,055 | 9.38 | 0.73 | 7.65 | 8.89 | 9.71 | 12.38 |
| $NO_2$ | 2,055 | 21.64 | 1.70 | 15.94 | 20.69 | 22.43 | 28.11 |
| NO | 2,055 | 21.39 | 2.45 | 14.26 | 20.04 | 22.47 | 31.31 |
| Per Capita Income | 2,051 | 35,196.41 | 27,024.55 | 2,634.11 | 20,610.11 | 38,194.48 | 222,941.30 |
| Change in 'Home' Devices (March 8 to May 11) | 2,053 | −0.14 | 0.17 | −0.90 | −0.20 | −0.05 | 0.61 |
| COVID-19 Deaths | 2,055 | 9.00 | 7.39 | 0 | 4 | 12 | 102 |
| COVID-19 Hospitalizations | 2,055 | 23.10 | 18.36 | 0 | 11 | 30 | 266 |
| COVID-19 Cases | 2,055 | 98.73 | 67.32 | 0 | 53 | 126 | 1,094 |

##### B. Outer Borough - 0.05-1km from Nearest Highway with Population > 500

| Statistic | N | Mean | St. Dev. | Min | Pctl(25) | Pctl(75) | Max |
|---|---|---|---|---|---|---|---|
| Tract Population | 1,254 | 3,976.86 | 2,159.08 | 514 | 2,401.5 | 5,082.8 | 28,272 |
| Downwind 3-5km (%) | 1,254 | 0.75 | 0.16 | 0.00 | 0.65 | 0.88 | 0.92 |
| Downwind 2-3km (%) | 1,254 | 0.61 | 0.19 | 0.00 | 0.48 | 0.76 | 0.92 |
| Downwind 1-2km (%) | 1,254 | 0.51 | 0.18 | 0.00 | 0.38 | 0.64 | 0.92 |
| Downwind 0.5-1km (%) | 1,254 | 0.26 | 0.16 | 0.00 | 0.15 | 0.35 | 0.81 |
| Downwind $<$ 0.5km (%) | 1,254 | 0.14 | 0.15 | 0 | 0 | 0.2 | 1 |
| Distance to Closest Highway | 1,254 | 0.46 | 0.26 | 0.05 | 0.23 | 0.68 | 1.00 |
| $PM_{2.5}$ | 1,254 | 9.29 | 0.54 | 7.91 | 8.86 | 9.69 | 10.88 |
| $NO_2$ | 1,254 | 21.42 | 1.22 | 16.67 | 20.66 | 22.19 | 24.91 |
| NO | 1,254 | 21.10 | 1.84 | 15.34 | 19.89 | 22.14 | 27.57 |
| Per Capita Income | 1,251 | 29,373.52 | 15,096.41 | 2,634.11 | 19,245.49 | 35,363.32 | 112,472.40 |
| Change in 'Home' Devices (March 8 to May 11) | 1,252 | −0.12 | 0.15 | −0.82 | −0.18 | −0.04 | 0.61 |
| COVID-19 Deaths | 1,254 | 9.69 | 8.10 | 0 | 4 | 13 | 102 |
| COVID-19 Hospitalizations | 1,254 | 25.28 | 20.76 | 0 | 12 | 34 | 266 |
| COVID-19 Cases | 1,254 | 106.70 | 75.47 | 0 | 54 | 138 | 1,094 |

*Notes:* Summary statistics for the Citywide sample are based on the characteristics of all census tracts in New York City with values for all variables. The Outer Borough sample includes census tracts in New York City outside of Manhattan below 110th Street and the statistics here are presented for a subset of these tracts with at least 500 residents that lie between 0.05 and 1km of the nearest highway.

### 2 Methods

It is well documented that air pollution is correlated with many socioeconomic variables that may influence COVID-19 outcomes. Even a rich set of control variables may result in biased estimates of the effect of air quality if some of the covariates have multi-directional causality with air quality, or if there are interaction effects between variables. To overcome this issue, we use an instrumental variables approach—relying on variations in pollution resulting from wind direction relative to nearby highways—to identify the causal effects of chronic ambient air pollution on COVID-19 disease intensity. A key assumption underlying this method is that wind direction is 'exogenous' or uncorrelated with individual or community characteristics that are correlated with COVID-19 outcomes. In other words, wind direction only effects COVID-19 outcomes through its effect on air quality.

The estimation is decomposed into two stages. In the first stage, we construct the instrumental variable $\widehat{AQ}$ by modelling $AQ$ as a linear combination of wind-related variables and other exogenous observables. $\widehat{AQ}$ can be considered the 'exogenous portion' of air quality. In the second stage, we regress COVID-19 outcomes on the exogenously measured $\widehat{AQ}$ and the same set of exogenous observables. This method is also called two stage least squares (2SLS) and is described in more detail below.

Section 2.1 describes how wind-related variables are constructed, section 2.2 provides the details on the first-stage regression, and section 2.3 provides the details on the second-stage regression.

#### 2.1 Constructing Wind-Related Instruments

Ambient pollutant concentrations will depend on both distance and direction to nearby highways. So, for each census tract $m$ we first calculate the angle $\theta_{mn}$ between the tract centroid and the nearest point on each highway segment $n$ within 5km of the tract centroid, as well as the distance $d_{mn}$ to the nearest point on each highway segment. A tract is considered downwind at a certain distance in any given hour if there exists at least one $\theta_{mn}$ such that the absolute difference between $\theta_{mn}$ and the wind direction measured at the nearest NCEI weather station is less than 45° and $d_{mn}$ lies within a certain range. The distance ranges that we use are: less than 0.5 km, 0.5-1km, 1 km-3km,

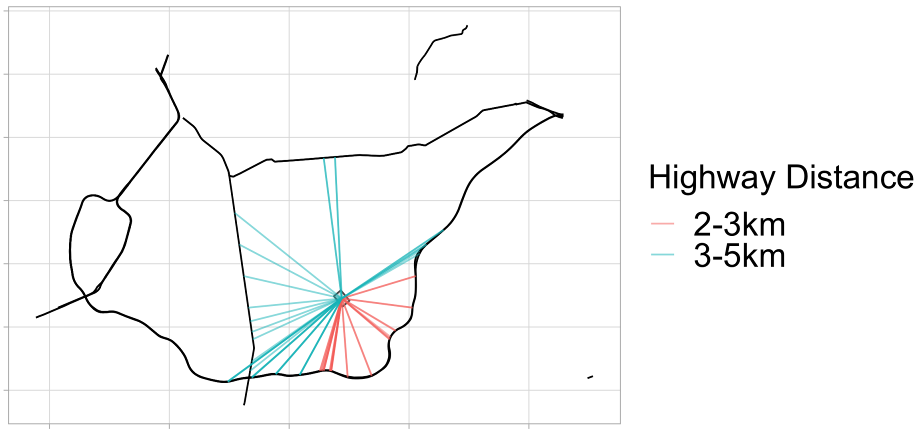

Figure S4: Calculating Distance and Orientation from a Census Tract to Nearby Highways

Note: Black lines show highways, red and blue rays show line from census tract centroid to closest point on each highway segment within 5km. Blue lines connect the tract to segments between 3 and 5km. Red lines go to segments between 2 and 3km. There are no road segments within 2km of this tract.

and 3-5km. We then calculate the fraction of hours during our 10-year period that each tract is downwind at each of these respective distance ranges. Next, we construct a set of corresponding instruments, denoted “$downwind_k$”, as the percent of time downwind of any highway segment that is located in range “k” km across a 10-year period.

Figure S4 illustrates this for an example census tract. The lines go from an example census tract centroid to the nearest point on each highway segment, with colors corresponding to distance groupings. This tract has no highway segment closer than 2km so the fraction of time downwind is 0 at those distances. This tract would be downwind at 5km for an hour where the wind blows from the North or West for example, and downwind at 3km for an hour where the wind blows from the South or East. Note, the tract could be downwind at multiple distances simultaneously. There is only 1 tract in NYC that has no roads within 5km.

It is critical to note that tracts that straddle both sides of a highway are subject to significant measurement error, as these tracts could be recorded as downwind (or not) on a day when a significant fraction of their population is actually upwind (or downwind). This measurement error is crucial, because tracts that are very close to highways are likely to differ in several respects compared to other tracts. For this reason, we drop tracts with centroids that are very close to a highway (within 50m). We also perform a robustness check that drops all tracts within 200m of a highway (see Table S14, panel B). For reference, the median area of an NYC census tract is slightly less than $2km^2$, or, if a square is assumed, 1.4km on a side. Figure S5 shows the fraction of time downwind at 1km, corresponding to our main specification, and 2km. Although there are geographic patterns in these variables, our identification relies on variation *within neighborhoods* (outlined in grey). Therefore, we essentially compare tracts that are on different sides of the same highway.

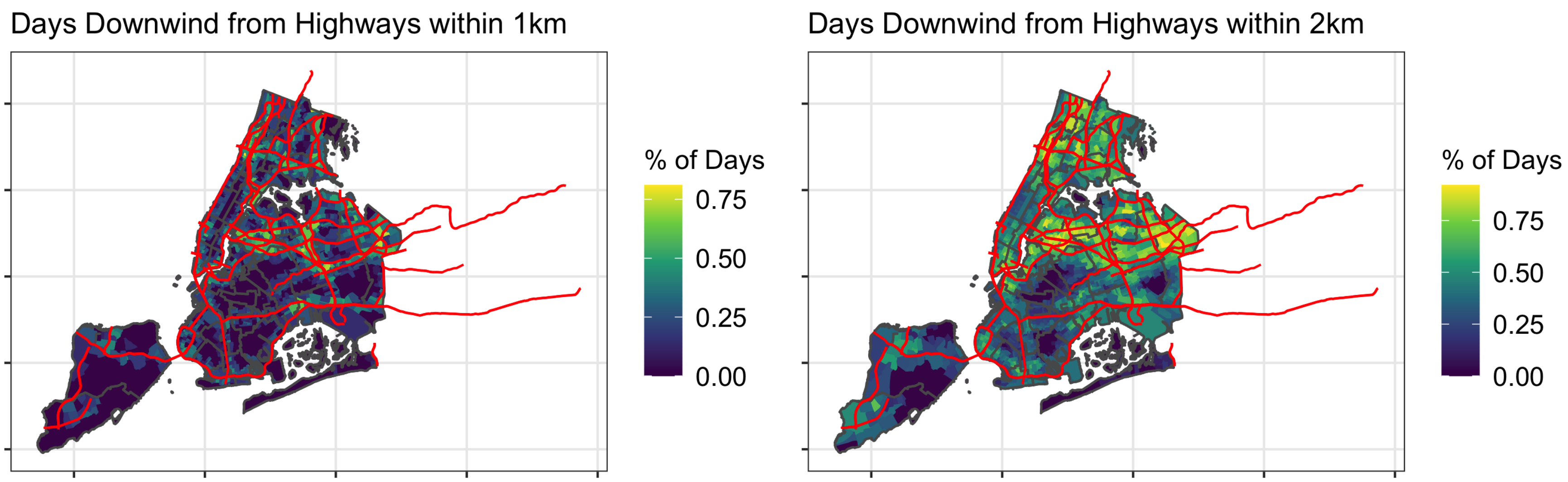

Figure S5: Fraction of time downwind for tracts within 1km and 2km.

Note: Highways in red, PUMAs outlined in grey.

#### 2.2 First-Stage Regression

In the first stage, we predict $PM_{2.5}$, $NO_2$ and NO as linear functions of ten-year average wind speed and the percent of time downwind of a highway at each distance:

$$\begin{aligned} AQ_i = &\eta + \sum_{k=1}^{K} \beta_k Downwind_{k,i} + \lambda Speed_i + \mu Distance_i \\ &+ PUMA_i + Station_i + \varepsilon_i \end{aligned} \tag{1}$$

$Downwind_{k,i}$ is the percent of time for census tract $i$ to be downwind of any highway segment that is located in the "$k$" km range across a 10 year period. For example, in models that we estimate on tracts within 1 km of a highway, we use two downwind variables: downwind at less than 0.5 km and between 0.5-1 km. The model also controls for 10-year average wind speed, *Speed*, distance to the closest highway, *Distance*, dummy variables for Public-Use-Microdata-Areas (PUMAs) and dummy variables for the nearest weather station. Wind speed is an important factor in pollutant dispersal, and adds significant predictive power to our first stage regressions. However, we also run a robustness check where we drop windspeed from our list of instruments (See Table S14E). We find nearly identical point estimates, but wider standard errors.

The key assumptions necessary for instrumental validity are (1) instruments are correlated with the pollutant concentrations (relevancy), and (2) instruments must not be related to COVID-19 disease intensity except through their relationships with pollutant concentrations (exclusion restriction). The relevancy assumption is met - as shown in Table S2 , the downwind variables are significantly associated with increased pollution concentration at distances less than 1 km. Focusing on the Outer Borough models, model (2) (5) and (8) suggest that a tract within .5km of a highway and downwind 100% of the time would have increased average ambient concentrations by 0.16 $\mu g/m^3$ of $PM_{2.5}$, 0.38 ppb of $NO_2$ and 0.73 ppb of NO, relative to a tract that is downwind 0% of the time. Our instruments perform notably worse in Manhattan due to more local traffic and other sources of pollutants, therefore we focus on results from our Outer Borough samples. Our weak instrument tests show F statistics greater than the traditional threshold of 10 for PM2.5 and $NO_2$, but not NO in the Outer Borough sample (see Table S4, panel A). Therefore, we also calculate weak-instrument robust critical-likelihood ratio confidence intervals

(Moriera 2003), shown in Table S4 in the 'Robust CI'. Our results remain significant at the 5% level.

#### Table S2: Results from First Stage Regressions

| | $PM_{2.5}$ | $PM_{2.5}$ | $PM_{2.5}$ | $NO_2$ | $NO_2$ | $NO_2$ | NO | NO | NO |
|---|---|---|---|---|---|---|---|---|---|
| | (1) | (2) | (3) | (4) | (5) | (6) | (7) | (8) | (9) |
| Downwind (3-5 km) | $-0.26^{***}$ | | | $-0.58^{***}$ | | | $-0.77^{***}$ | | |
| | (0.06) | | | (0.14) | | | (0.24) | | |
| Downwind (2-3 km) | 0.04 | | | 0.04 | | | 0.01 | | |
| | (0.04) | | | (0.09) | | | (0.16) | | |
| Downwind (1-2 km) | $0.07^{**}$ | | | $0.17^{**}$ | | | 0.22 | | |
| | (0.03) | | | (0.07) | | | (0.14) | | |
| Downwind (0.5-1 km) | $0.09^{***}$ | $0.12^{***}$ | 0.23 | $0.20^{***}$ | $0.21^{***}$ | $0.96^{*}$ | $0.45^{***}$ | $0.40^{***}$ | $1.86^{**}$ |
| | (0.03) | (0.03) | (0.21) | (0.07) | (0.07) | (0.50) | (0.14) | (0.14) | (0.92) |
| Downwind (less than 0.5 km) | $0.10^{**}$ | $0.16^{***}$ | $-0.19$ | $0.31^{***}$ | $0.38^{***}$ | $-0.11$ | $0.66^{***}$ | $0.73^{***}$ | $-0.12$ |
| | (0.04) | (0.05) | (0.32) | (0.10) | (0.11) | (0.77) | (0.19) | (0.23) | (1.46) |
| Wind speed | $-0.75^{***}$ | $-0.59^{***}$ | $-4.84^{***}$ | $-1.28^{***}$ | $-1.11^{***}$ | $-10.73^{***}$ | $-1.81^{***}$ | $-1.65^{***}$ | $-19.49^{***}$ |
| | (0.13) | (0.14) | (0.94) | (0.29) | (0.31) | (2.33) | (0.55) | (0.58) | (4.20) |
| Constant | $12.81^{***}$ | $12.01^{***}$ | $29.40^{***}$ | $26.98^{***}$ | $25.91^{***}$ | $65.64^{***}$ | $29.29^{***}$ | $28.21^{***}$ | $99.24^{***}$ |
| | (0.56) | (0.57) | (3.96) | (1.29) | (1.30) | (9.84) | (2.41) | (2.41) | (17.76) |
| Geography | Citywide | Outer Borough | Manhattan | Citywide | Outer Borough | Manhattan | Citywide | Outer Borough | Manhattan |
| Maximum Distance from Highway | All | 1 km | 1 km | All | 1 km | 1 km | All | 1 km | 1 km |
| $N$ | 2,055 | 1,254 | 153 | 2,055 | 1,254 | 153 | 2,055 | 1,254 | 153 |

*Notes:* The dependent variable in these OLS models is the inverse-distance weighted chronic ambient concentration of each pollutant at the centroid of the census tracts included in each sample. The reported coefficients are based on samples that include all tracts within the given Geography at least 50 meters from the nearest highway with more than 500 people. Each model includes the following additional controls: distance to the nearest highway, and indicators for PUMA and the nearest weather station. Robust standard errors are reported in parentheses. One, two, and three stars indicate 10 percent, 5 percent, and 1 percent significance, respectively.

As for the exclusion restriction assumption, while exposure to poor air quality as a result of living near a highway is likely endogenous with factors related to COVID-19 disease intensity (e.g., income, health care access, etc.), we assume that, conditional on living near a highway and in the same neighborhood, our instruments only affect COVID-19 outcomes through their effect on pollution concentration. This is a reasonable assumption because the pollutants of interest are generally not detectable via sight or smell at concentrations in NYC, and differences of the magnitudes our coefficients report would clearly not be detectable. We also conduct Sargan's over-identification tests, which have p-values greater than .05 in all of our Outer Borough 1km specifications, as well as most other specifications (see Table S4, panel A).

We further test the assumption that tracts within the same neighborhood at the same distance from a highway that are either primarily upwind or downwind of highways are comparable by conducting balance checks. Table S3 presents the results of these tests for tracts within 0.5km and 1km of a highway. In these balance checks, we use the mean fraction of time tracts within these distances are downwind to define which tracts are upwind or downwind of a highway. A tract is considered primarily downwind of a highway if its time spent downwind is greater than the mean time spent downwind, and is considered upwind otherwise. The balance checks for tracts within 0.5km and 1km of a highway do not show any variables with a normalized difference between upwind and downwind groups greater than 0.25. This supports our assumption that conditional on living near a highway, there are no significant socioeconomic or demographic differences between tracts that are primarily upwind or downwind of a highway. While there are a number of reasons for residential sorting to occur in different areas of NYC, based on our analysis of the tracts closest to highways and the fact that there is not a prevailing wind direction in NYC, we believe it is unlikely that individuals are sorting based on time spent upwind or downwind of a highway. For these reasons, we assume that our instruments are exogenous and that the exclusion restriction is met.

### Table S3: Balance Checks

##### A. Balance Check for Census Tracts within .5km of a Highway

| 5-Year 2018 ACS Variable | Downwind | Upwind | Normalized Difference |
|---|---|---|---|
| Percentage White | 0.317 | 0.335 | -0.062 |
| | [0.010] | [0.027] | |
| Percentage Black | 0.179 | 0.172 | 0.029 |
| | [0.009] | [0.021] | |
| Percentage Asian | 0.165 | 0.129 | 0.196 |
| | [0.007] | [0.014] | |
| Percentage Latinx | 0.306 | 0.330 | -0.100 |
| | [0.008] | [0.023] | |
| Percentage Non-Latinx Other Race | 0.032 | 0.034 | -0.035 |
| | [0.001] | [0.004] | |
| 2018 Tract population | 4021.729 | 4545.057 | -0.218 |
| | [82.788] | [255.041] | |
| Percentage Safegraph Device Change | -0.069 | -0.102 | 0.186 |
| | [0.006] | [0.014] | |
| Percentage Per-Capita Income ¡ $10,000 | 0.085 | 0.089 | -0.057 |
| | [0.002] | [0.005] | |
| Gini coefficient | 0.461 | 0.473 | -0.198 |
| | [0.002] | [0.006] | |
| Percentage Aged Under 17 | 0.152 | 0.151 | 0.023 |
| | [0.002] | [0.005] | |
| Percentage Aged 18-24 | 0.646 | 0.643 | 0.043 |
| | [0.002] | [0.005] | |
| Percentage Aged 45-54 | 0.073 | 0.075 | -0.066 |
| | [0.001] | [0.003] | |
| Percentage Aged 55-64 | 0.063 | 0.063 | -0.008 |
| | [0.001] | [0.002] | |
| Percentage Aged 65-74 | 0.057 | 0.058 | -0.020 |
| | [0.001] | [0.002] | |
| Percentage Aged 75 and up | 0.089 | 0.089 | 0.001 |
| | [0.001] | [0.002] | |
| Percentage Uninsured | 0.086 | 0.082 | 0.059 |
| | [0.002] | [0.004] | |
| Percentage with Bachelor's degree | 0.213 | 0.207 | 0.058 |
| | [0.004] | [0.011] | |
| Percentage with Advanced degree | 0.147 | 0.139 | 0.073 |
| | [0.004] | [0.010] | |
| Percentage Employed in Service Industry | 0.234 | 0.235 | -0.005 |
| | [0.004] | [0.010] | |
| Percentage Employed in Production Industry | 0.101 | 0.096 | 0.082 |
| | [0.002] | [0.005] | |
| Percentage spending at least 50% on Rent | 0.304 | 0.311 | -0.059 |
| | [0.004] | [0.011] | |

*Notes:* This table is a balance check of control variables from the 5-year 2018 ACS at the census tract level. Each census tract within 0.5km of a highway was given a 'downwind' or 'upwind' designation. In this balance check, a census tract is considered to be 'downwind' if the amount of time a tract is downwind of a highway is greater than the mean time spent downwind for all tracts within 0.5km from 2008-2018, and is considered 'upwind' otherwise.

### Table S3: Balance Checks (continued)

**B. Balance Check for Census Tracts within 1km of a Highway**

| 5-Year 2018 ACS Variable | Downwind | Upwind | Normalized Difference |
|---|---|---|---|
| Percentage White | 0.321 | 0.338 | -0.057 |
| | [0.009] | [0.014] | |
| Percentage Black | 0.175 | 0.221 | -0.180 |
| | [0.008] | [0.013] | |
| Percentage Asian | 0.163 | 0.141 | 0.127 |
| | [0.006] | [0.008] | |
| Percentage Latinx | 0.309 | 0.266 | 0.182 |
| | [0.008] | [0.010] | |
| Percentage Non-Latinx Other Race | 0.032 | 0.035 | -0.076 |
| | [0.001] | [0.002] | |
| 2018 Tract population | 4242.453 | 3724.026 | 0.212 |
| | [80.058] | [98.484] | |
| Percentage Safegraph Device Change | -0.077 | -0.079 | 0.008 |
| | [0.006] | [0.011] | |
| Percentage Per-Capita Income ¡ $10,000 | 0.085 | 0.084 | 0.016 |
| | [0.002] | [0.003] | |
| Gini coefficient | 0.463 | 0.463 | 0.002 |
| | [0.002] | [0.003] | |
| Percentage Aged Under 17 | 0.149 | 0.155 | -0.141 |
| | [0.001] | [0.002] | |
| Percentage Aged 18-24 | 0.646 | 0.646 | -0.014 |
| | [0.002] | [0.003] | |
| Percentage Aged 45-54 | 0.074 | 0.072 | 0.081 |
| | [0.001] | [0.001] | |
| Percentage Aged 55-64 | 0.063 | 0.061 | 0.099 |
| | [0.001] | [0.001] | |
| Percentage Aged 65-74 | 0.058 | 0.058 | -0.006 |
| | [0.001] | [0.001] | |
| Percentage Aged 75 and up | 0.088 | 0.091 | -0.085 |
| | [0.001] | [0.001] | |
| Percentage Uninsured | 0.086 | 0.083 | 0.049 |
| | [0.002] | [0.002] | |
| Percentage with Bachelor's degree | 0.216 | 0.208 | 0.082 |
| | [0.004] | [0.005] | |
| Percentage with Advanced degree | 0.152 | 0.142 | 0.078 |
| | [0.004] | [0.005] | |
| Percentage Employed in Service Industry | 0.232 | 0.239 | -0.059 |
| | [0.004] | [0.005] | |
| Percentage Employed in Production Industry | 0.099 | 0.098 | 0.022 |
| | [0.002] | [0.003] | |
| Percentage spending at least 50% on Rent | 0.301 | 0.315 | -0.117 |
| | [0.004] | [0.006] | |

*Notes:* This table is a balance check of control variables from the 5-year 2018 ACS at the census tract level. Each census tract within 1km of a highway was given a 'downwind' or 'upwind' designation. In this balance check, a census tract is considered to be 'downwind' if the amount of time a tract is downwind of a highway is greater than the mean time spent downwind for all tracts within 1km from 2008-2018, and is considered 'upwind' otherwise.

#### 2.3 Second-Stage Regression

##### 2.3.1 Log-linear model

Our second stage regressions estimate the causal impact of a change in ambient pollutant concentration on two different measures of the intensity of COVID-19 disease: deaths and hospitalizations. Given the discovery that rate-based measures of these outcomes, used to control for different populations across tracts, are subject to systematic measurement error, which would lead to biased estimates in our regressions, we leave our dependent variables as counts. Thus our results can be interpreted as the change in deaths in an average tract, as our instruments are exogenous to tract population. Our key specification is:

$$\log(Y_i + 1) = \alpha_0 + \alpha_1 \widehat{AQ}_i + \alpha_2 Distance_i + PUMA_i + Station_i + \nu_i \tag{2}$$

$Y_i$ are counts of deaths and hospitalizations. We use log-transformed counts and add one to the count variables to adjust for tracts with zero deaths or hospitalizations. $\widehat{AQ}_i$ is the instrumented measure of ambient concentration for $PM_{2.5}$, $NO_2$, and NO predicted from equation (1). We include weather station and PUMA dummy variables in the second stage as well. The key parameter of interest is $\alpha_1$. Our identification strategy compares tracts within the same PUMA that lie within the same distance of the highway that spent different amounts of time downwind from 2009-2018. We exclude census tracts with centroids located within 50m of the closest highway and with populations smaller than 500 as discussed above. There are 2055 census tracts in NYC located in 55 PUMAs that fit this criteria.

We define three geographies for use in our analyses: *Citywide*, including all tracts across New York City, *Outer Borough*, including all tracts in Brooklyn, the Bronx, Queens, and Staten Island, in addition to all tracts above 110th Street in Manhattan, and *Manhattan*, which actually only includes Manhattan below 110th Street. The initial review of our Safegraph and ACS data indicated that census tracts below 110th in Manhattan differ systematically from the rest of NYC, and we find different effects of pollution on COVID-19 between these two geographies.

##### 2.3.2 Poisson model

We also estimate a Poisson model using an instrumental variable control function estimator proposed by Wooldridge (2010) to avoid the incidental parameters problem from the large number of PUMA dummy variables. This method uses the residuals $\hat{\epsilon}_i$ estimated from equation (1) in the second stage:

$$Y_i = \mu_i \exp(\gamma_1 AQ_i + \gamma_2 \hat{\epsilon}_i + \gamma_3 Distance_i + Station_i) \exp(\nu_i) \quad (3)$$

In the control function approach, the residuals, $\hat{\epsilon}_i$, control for the endogenous portion of air quality. The PUMA dummies, $\mu_i$ are multiplicative. We estimate the coefficients using maximum likelihood.

### 3 Results and Discussions

#### 3.1 Main Specifications

Our main specifications, presented in panel A of Tables S4 and S5, are estimated with equation (2) using tracts with centroids that lie within 0.05-1km of a highway and population greater than 500. Table S4 reports the coefficient estimates for our focal pollutants where the outcome variables are log transformed death counts. Table S5 reports the coefficient estimates for the log transformed number of hospitalizations as the outcome.

From (2), we can estimate the marginal increase in death or hospitalization counts associated with an increase in the 10-year average pollutant concentration. Given large coefficients, typical approximations to percent increases in log-transformed variables are inaccurate for our models. Also, given the limited variation in pollution explained by our instruments, we calculate Average Marginal Effects (AMEs) corresponding to an increase commensurate with a tract going from downwind of a highway within .5 km 0% of the time to 100% of the time. This amounts to an increase of 0.16 $\mu g/m^3$ of $PM_{2.5}$, 0.38 ppb of $NO_2$ and 0.73 ppb of NO. We bootstrap 95% confidence intervals for AMEs using 1000 replicates. These estimates and confidence intervals are reported in Tables S4, S5, S8 and S9.

#### Table S4: Effects on Deaths

**A. Deaths - log-linear (main specifications)**

| | (1) | (2) | (3) | (4) | (5) | (6) | (7) | (8) | (9) |
|---|---|---|---|---|---|---|---|---|---|
| | $PM_{2.5}$ | $PM_{2.5}$ | $PM_{2.5}$ | $NO_2$ | $NO_2$ | $NO_2$ | NO | NO | NO |
| AQ | $0.91^{**}$ | $1.39^{**}$ | $-0.20$ | $0.43^{**}$ | $0.66^{**}$ | $-0.10$ | $0.25^{**}$ | $0.36^{**}$ | $-0.06$ |
| | (0.45) | (0.61) | (0.29) | (0.22) | (0.31) | (0.13) | (0.13) | (0.19) | (0.07) |
| Robust CIs | (0.1, 1.95) | (0.28, 2.84) | (-0.9, 0.59) | (0.04, 0.96) | (0.1, 1.45) | (-0.42, 0.26) | (0.02, 0.58) | (0.04, 0.9) | (-0.24, 0.14) |
| AME | 1.73 | 2.83 | -0.27 | 1.96 | 3.25 | -0.31 | 2.23 | 3.54 | -0.33 |
| | (0.13, 4.64) | (0.43, 8.11) | (-1.13, 0.64) | (0.23, 5.46) | (0.21, 8.3) | (-1.22, 0.69) | (-0.03, 5.71) | (0.07, 11.35) | (-1.29, 0.58) |
| Geography | Citywide | Outer Boroughs | Manhattan | Citywide | Outer Boroughs | Manhattan | Citywide | Outer Boroughs | Manhattan |
| F statistic | 18.49 | 13.74 | 9.36 | 16.12 | 11.1 | 8.4 | 12.98 | 7.85 | 8.82 |
| Sargan p value | 0.48 | 0.49 | 0.61 | 0.43 | 0.34 | 0.64 | 0.44 | 0.28 | 0.64 |
| Adjusted $R^2$ | 0.13 | 0.12 | 0.22 | 0.12 | 0.11 | 0.22 | 0.09 | 0.08 | 0.22 |
| *N* | 1,407 | 1,254 | 153 | 1,407 | 1,254 | 153 | 1,407 | 1,254 | 153 |

**B. Deaths - Poisson**

| | (1) | (2) | (3) | (4) | (5) | (6) | (7) | (8) | (9) |
|---|---|---|---|---|---|---|---|---|---|
| | $PM_{2.5}$ | $PM_{2.5}$ | $PM_{2.5}$ | $NO_2$ | $NO_2$ | $NO_2$ | NO | NO | NO |
| AQ | 1.47 | 1.98 | $-0.43$ | 0.68 | 0.92 | $-0.20$ | 0.38 | 0.49 | $-0.11$ |
| | (0.31, 2.86) | (0.44, 3.75) | ($-1.06$, 0.36) | (0.12, 1.31) | (0.16, 1.99) | ($-0.46$, 0.12) | (0.05, 0.74) | (0.03, 1.02) | ($-0.26$, 0.07) |
| AME | 2.49 | 3.62 | -0.49 | 2.76 | 4.04 | -0.52 | 3.05 | 4.16 | -0.54 |
| | (0.37, 5.75) | (0.67, 8.25) | (-1.12, 0.35) | (0.24, 5.91) | (0.29, 9.48) | (-1.18, 0.35) | (0.4, 7.18) | (0.16, 12.4) | (-1.28, 0.43) |
| Geography | Citywide | Outer Boroughs | Manhattan | Citywide | Outer Boroughs | Manhattan | Citywide | Outer Boroughs | Manhattan |
| *N* | 1,407 | 1,254 | 153 | 1,407 | 1,254 | 153 | 1,407 | 1,254 | 153 |

**C. Deaths - OLS**

| | (1) | (2) | (3) | (4) | (5) | (6) | (7) | (8) | (9) |
|---|---|---|---|---|---|---|---|---|---|
| | $PM_{2.5}$ | $PM_{2.5}$ | $PM_{2.5}$ | $NO_2$ | $NO_2$ | $NO_2$ | NO | NO | NO |
| AQ | $-0.25^{**}$ | $-0.16$ | $-0.40^{***}$ | $-0.10^{**}$ | $-0.05$ | $-0.16^{***}$ | $-0.06^{**}$ | $-0.04$ | $-0.09^{***}$ |
| | (0.10) | (0.13) | (0.14) | (0.04) | (0.06) | (0.06) | (0.02) | (0.03) | (0.03) |
| Geography | Citywide | Outer Boroughs | Manhattan | Citywide | Outer Boroughs | Manhattan | Citywide | Outer Boroughs | Manhattan |
| Adjusted $R^2$ | 0.23 | 0.22 | 0.23 | 0.23 | 0.22 | 0.23 | 0.23 | 0.23 | 0.22 |
| *N* | 1,407 | 1,254 | 153 | 1,407 | 1,254 | 153 | 1,407 | 1,254 | 153 |

*Notes:* The dependent variable in the 2SLS log-linear models (main specifications) and OLS models is the logged count plus one of deaths measured at the census tract level, while it is simply the count of deaths in the 2SLS Poisson models. The reported coefficients are based on samples that include all tracts within the given Geography that include at least 500 people, according to the 2018 5-year ACS, with centroids that lie between 0.05 and 1km from the nearest highway segment. Each model includes the following additional controls: distance to the nearest highway, and indicators for PUMA and nearest weather station. Robust standard errors are reported in parentheses, except for the Poisson models which report 95% bootstrapped confidence intervals. One, two, and three stars indicate 10 percent, 5 percent, and 1 percent significance in the OLS and log-linear panels, respectively. Average Marginal Effects (AME) are also reported for log-linear and Poisson models with 95% bootstrapped confidence intervals. Weak-instrument robust confidence intervals (95%) based on the critical-likelihood ratio test are reported for log-linear models.

#### Table S5: Effects on Hospitalizations

**A. Hospitalizations - log-linear (main specifications)**

| | (1) | (2) | (3) | (4) | (5) | (6) | (7) | (8) | (9) |
|---|---|---|---|---|---|---|---|---|---|
| | $PM_{2.5}$ | $PM_{2.5}$ | $PM_{2.5}$ | $NO_2$ | $NO_2$ | $NO_2$ | NO | NO | NO |
| AQ | 0.56 | $1.11^{*}$ | $-0.40$ | 0.27 | $0.50^{*}$ | $-0.17$ | 0.17 | $0.30^{*}$ | $-0.09$ |
| | (0.45) | (0.60) | (0.31) | (0.21) | (0.30) | (0.14) | (0.12) | (0.18) | (0.08) |
| AME | 2.45 | 5.33 | -1.26 | 2.81 | 5.81 | -1.25 | 3.52 | 6.79 | -1.3 |
| | (-1.46, 8.16) | (-0.13, 16.04) | (-3.3, 1.44) | (-1.38, 11.38) | (-0.73, 18.74) | (-3.72, 1.74) | (-1.53, 12.09) | (-1.04, 26.6) | (-3.69, 1.87) |
| Geography | Citywide | Outer Boroughs | Manhattan | Citywide | Outer Boroughs | Manhattan | Citywide | Outer Boroughs | Manhattan |
| F statistic | 18.49 | 13.74 | 9.36 | 16.12 | 11.1 | 8.4 | 12.98 | 7.85 | 8.82 |
| Sargan p value | 0.48 | 0.49 | 0.61 | 0.43 | 0.34 | 0.64 | 0.44 | 0.28 | 0.64 |
| Adjusted $R^2$ | 0.26 | 0.24 | 0.20 | 0.26 | 0.25 | 0.20 | 0.23 | 0.21 | 0.19 |
| $N$ | 1,407 | 1,254 | 153 | 1,407 | 1,254 | 153 | 1,407 | 1,254 | 153 |

**B. Hospitalizations - Poisson**

| | (1) | (2) | (3) | (4) | (5) | (6) | (7) | (8) | (9) |
|---|---|---|---|---|---|---|---|---|---|
| | $PM_{2.5}$ | $PM_{2.5}$ | $PM_{2.5}$ | $NO_2$ | $NO_2$ | $NO_2$ | NO | NO | NO |
| AQ | 1.02 | 1.61 | $-0.50$ | 0.48 | 0.74 | $-0.21$ | 0.29 | 0.42 | $-0.11$ |
| | $(-0.16, 2.30)$ | (0.07, 3.39) | $(-0.98, 0.25)$ | $(-0.07, 1.04)$ | (0.03, 1.57) | $(-0.44, 0.12)$ | $(-0.02, 0.64)$ | $(-0.01, 0.94)$ | $(-0.24, 0.07)$ |
| AME | 4.36 | 7.4 | -1.47 | 4.96 | 8.22 | -1.49 | 5.82 | 9.15 | -1.52 |
| | (-0.58, 11.46) | (0.82, 18.12) | (-3.07, 0.66) | (-0.5, 13.13) | (0.54, 21.88) | (-2.99, 0.8) | (-0.1, 15.51) | (0.74, 27.31) | (-3.33, 1.04) |
| Geography | Citywide | Outer Boroughs | Manhattan | Citywide | Outer Boroughs | Manhattan | Citywide | Outer Boroughs | Manhattan |
| $N$ | 1,407 | 1,254 | 153 | 1,407 | 1,254 | 153 | 1,407 | 1,254 | 153 |

**C. Hospitalizations - OLS**

| | (1) | (2) | (3) | (4) | (5) | (6) | (7) | (8) | (9) |
|---|---|---|---|---|---|---|---|---|---|
| | $PM_{2.5}$ | $PM_{2.5}$ | $PM_{2.5}$ | $NO_2$ | $NO_2$ | $NO_2$ | NO | NO | NO |
| AQ | $-0.29^{***}$ | $-0.25^{**}$ | $-0.38^{***}$ | $-0.11^{***}$ | $-0.08$ | $-0.15^{***}$ | $-0.07^{***}$ | $-0.07^{**}$ | $-0.08^{***}$ |
| | (0.09) | (0.13) | (0.13) | (0.04) | (0.06) | (0.05) | (0.02) | (0.03) | (0.03) |
| Geography | Citywide | Outer Boroughs | Manhattan | Citywide | Outer Boroughs | Manhattan | Citywide | Outer Boroughs | Manhattan |
| Adjusted $R^2$ | 0.31 | 0.32 | 0.20 | 0.31 | 0.32 | 0.20 | 0.31 | 0.33 | 0.19 |
| $N$ | 1,407 | 1,254 | 153 | 1,407 | 1,254 | 153 | 1,407 | 1,254 | 153 |

*Notes:* The dependent variable in the 2SLS log-linear models (main specifications) and OLS models is the logged count plus one of hospitalizations measured at the census tract level, while it is simply the count of hospitalizations in the Poisson models. The reported coefficients are based on samples that include all tracts within the given Geography that include at least 500 people, according to the 2018 5-year ACS, with centroids that lie between 0.05 and 1km from the nearest highway segment. Each model includes the following additional controls: distance to the nearest highway, and indicators for PUMA and nearest weather station. Robust standard errors are reported in parentheses, except for the Poisson models which report 95% bootstrapped confidence intervals. One, two, and three stars indicate 10 percent, 5 percent, and 1 percent significance in the OLS and log-linear panels, respectively. Average Marginal Effects (AME) are also reported for log-linear and Poisson models with 95% bootstrapped confidence intervals.

Tables S4 and S5, panel B report the results estimated with Poisson models as specified in equation (3). 95% confidence intervals for coefficients and marginal effects in the Poisson model are bootstrapped with 1000 replications (Table S4 and S5). The marginal effects of an increase in ambient pollution associated with a tract being continuously downwind of a highway less than .5 km away are comparable but significantly higher than the log-transformed model: we find 7.4 more deaths compared to 5.33 in the Outer Borough sample for $PM_{2.5}$; 8.22 compared to 5.81 for $NO_2$, and 9.15 compared to 6.79 for NO.

We also report in Tables S4 and S5, panel C estimates from OLS models that do not account for the endogeneity of air pollution to contrast with the instrumental variable estimates in panel A and B.

#### 3.2 Predicted Deaths and Hospitalizations

We predict the number of deaths and hospitalizations at representative concentrations of each of the pollutants using both the log-linear (equation (2)) and Poisson model (equation (3)). These results are presented in Tables S6 and S7. We calculate 95% confidence intervals using 1000 bootstrapped replicates. In general, the models agree, although the confidence intervals in the Poisson regressions are wider at higher concentrations. The extremely wide confidence intervals at high concentrations can be explained by the paucity of census tracts with long-term pollutant concentrations at those levels in our sample.

#### Table S6: Predicted Deaths

| | | | | | | | | |
|---|---|---|---|---|---|---|---|---|
| | | | | **A. $PM_{2.5}$** | | | | |
| Concentration | 7 | 7.5 | 8 | 8.5 | 9 | 9.5 | 10 | 10.5 |
| log-linear | −0.45 | 0.09 | 1.19 | 3.38 | 7.78 | 16.59 | 34.23 | 69.58 |
| | (−0.95, 6.47) | (−0.84, 6.30) | (−0.27, 7.02) | (1.82, 8.09) | (7.38, 10.11) | (10.12, 39.98) | (11.95, 182.60) | (12.01, 764.61) |
| Poisson | 0.15 | 0.41 | 1.11 | 3.00 | 8.09 | 21.81 | 58.75 | 158.30 |
| | (0.01, 3.44) | (0.04, 4.21) | (0.32, 6.08) | (2.05, 6.88) | (7.41, 15.51) | (10.19, 110.20) | (11.89, 546.45) | (20.08, 4,161.36) |
| | | | | **B. $NO_2$** | | | | |
| Concentration | 16 | 17 | 18 | 19 | 20 | 21 | 22 | 23 |
| log-linear | −0.59 | −0.21 | 0.52 | 1.93 | 4.67 | 9.94 | 20.14 | 39.82 |
| | (−0.96, 7.49) | (−0.85, 7.92) | (−0.42, 8.05) | (0.93, 7.93) | (4.18, 9.94) | (8.61, 33.45) | (10.15, 131.42) | (10.19, 532.83) |
| Poisson | 0.13 | 0.32 | 0.81 | 2.03 | 5.08 | 12.72 | 31.85 | 79.75 |
| | (0.02, 3.83) | (0.10, 5.03) | (0.46, 5.57) | (1.65, 8.30) | (4.54, 21.19) | (8.59, 216.91) | (10.71, 1,270.25) | (11.67, 7,712.02) |
| | | | | **C. NO** | | | | |
| Concentration | 16 | 17 | 18 | 19 | 20 | 21 | 22 | 23 |
| log-linear | 1.00 | 1.88 | 3.15 | 4.97 | 7.60 | 11.39 | 16.84 | 24.69 |
| | (−0.48, 9.13) | (0.14, 9.63) | (1.40, 9.38) | (3.88, 9.70) | (7.24, 12.76) | (9.52, 23.44) | (9.55, 46.57) | (9.83, 109.34) |
| Poisson | 1.09 | 1.77 | 2.89 | 4.72 | 7.71 | 12.57 | 20.51 | 33.46 |
| | (0.23, 8.10) | (0.69, 7.86) | (1.82, 9.30) | (4.04, 9.36) | (7.21, 14.72) | (9.49, 48.15) | (10.07, 128.36) | (10.58, 361.81) |

*Notes:* These tables report the predicted deaths at various chronic ambient pollutant concentrations, based on the results of our 2SLS log-linear and Poisson models. The prediction is generated using the estimated coefficients and holding the concentration of the focal pollutant at the listed value across all tracts in our Outer Bourough sample that include at least 500 people, according to the 2018 5-year ACS, with centroids that lie between 0.05 and 1km meters from the nearest highway segment. The 95% confidence interval, based on a percentile bootstrap of 1,000 replications, is presented in parentheses.

Table S7: Predicted Hospitalizations

| | | | | | | | | |
|---|---|---|---|---|---|---|---|---|
| | | | | **A. $PM_{2.5}$** | | | | |
| Concentration | 7 | 7.5 | 8 | 8.5 | 9 | 9.5 | 10 | 10.5 |
| log-linear | 1.33 | 3.05 | 6.05 | 11.25 | 20.29 | 36.00 | 63.32 | 110.79 |
| | (−0.86, 29.04) | (−0.30, 26.74) | (1.02, 27.28) | (5.90, 24.84) | (19.39, 26.96) | (25.42, 91.01) | (23.14, 288.35) | (25.19, 887.60) |
| Poisson | 0.80 | 1.79 | 4.00 | 8.94 | 19.94 | 44.50 | 99.29 | 221.56 |
| | (0.03, 18.71) | (0.15, 20.80) | (1.09, 17.86) | (5.42, 21.10) | (19.05, 30.60) | (25.74, 158.94) | (29.24, 1,099.01) | (31.38, 4,658.52) |
| | | | | **B. $NO_2$** | | | | |
| Concentration | 16 | 17 | 18 | 19 | 20 | 21 | 22 | 23 |
| log-linear | 0.66 | 1.38 | 2.89 | 6.06 | 12.70 | 26.65 | 55.89 | 117.22 |
| | (0.06, 21.18) | (0.31, 23.28) | (1.30, 20.72) | (4.37, 25.35) | (11.83, 40.26) | (22.05, 202.53) | (25.76, 1,093.79) | (26.46, 5,261.68) |
| Poisson | 0.66 | 1.38 | 2.89 | 6.06 | 12.70 | 26.65 | 55.89 | 117.22 |
| | (0.06, 21.18) | (0.31, 23.28) | (1.30, 20.72) | (4.37, 25.35) | (11.83, 40.26) | (22.05, 202.53) | (25.76, 1,093.79) | (26.46, 5,261.68) |
| | | | | **C. NO** | | | | |
| Concentration | 16 | 17 | 18 | 19 | 20 | 21 | 22 | 23 |
| log-linear | 5.44 | 7.67 | 10.68 | 14.72 | 20.17 | 27.50 | 37.38 | 50.67 |
| | (0.24, 31.78) | (1.96, 31.33) | (5.06, 31.72) | (10.93, 28.53) | (19.20, 28.40) | (24.64, 57.47) | (24.56, 116.29) | (22.73, 208.73) |
| Poisson | 3.61 | 5.51 | 8.42 | 12.86 | 19.63 | 29.97 | 45.76 | 69.87 |
| | (0.76, 22.79) | (1.89, 24.79) | (5.09, 27.24) | (10.77, 26.98) | (18.70, 37.83) | (24.58, 103.48) | (25.50, 229.17) | (26.60, 870.86) |

*Notes:* These tables report the predicted hospitalizations at various chronic ambient pollutant concentrations, based on the results of our 2SLS log-linear and Poisson models. The prediction is generated using the estimated coefficients and holding the concentration of the focal pollutant at the listed value across all tracts in our Outer Bourough sample that include at least 500 people, according to the 2018 5-year ACS, with centroids that lie between 0.05 and 1km from the nearest highway segment. The 95% confidence interval, based on a percentile bootstrap of 1,000 replications, is presented in parentheses.

#### 3.3 Near Distance Analysis

As TRAP concentrations decay quickly with distance from highways (Karner et al. 2010), we explore whether the precision of our estimates increases as we restrict our observations to tracts within a certain distance of the highway. Within each geography, we run regressions for all tracts, and tracts within 2km, 1km, and .5km of a highway. These results are presented in Tables S8 and S9.

### Table S8: Near Distance Analysis on Deaths

| | | | | | | | | | | | | |
|---|---|---|---|---|---|---|---|---|---|---|---|---|
| | **A. $PM_{2.5}$** | | | | | | | | | | | |
| | All | 2km | 1km | .5km | All | 2km | 1km | .5km | All | 2km | 1km | .5km |
| | (1) | (2) | (3) | (4) | (5) | (6) | (7) | (8) | (9) | (10) | (11) | (12) |
| $PM_{2.5}$ | $0.84^{**}$ | $0.95^{**}$ | $0.91^{**}$ | $1.02^{*}$ | $0.85^{**}$ | $1.03^{**}$ | $1.39^{**}$ | $1.52^{**}$ | $-0.09$ | $-0.13$ | $-0.20$ | $-0.44$ |
| | (0.38) | (0.41) | (0.45) | (0.54) | (0.39) | (0.47) | (0.61) | (0.75) | (0.25) | (0.27) | (0.29) | (0.33) |
| AME | 1.48 | 1.75 | 1.73 | 1.99 | 1.53 | 1.92 | 2.83 | 3.17 | -0.11 | -0.16 | -0.27 | -0.61 |
| | (0.03, 3.28) | (0.21, 4.05) | (0.06, 4.61) | (-0.03, 6.09) | (0.04, 3.34) | (0.16, 4.51) | (0.44, 7.18) | (0.1, 9.63) | (-0.75, 0.55) | (-0.8, 0.59) | (-1.15, 0.65) | (-1.69, 0.71) |
| Geography | All | All | All | All | Outer-Borough | Outer-Borough | Outer-Borough | Outer-Borough | Manhattan | Manhattan | Manhattan | Manhattan |
| F statistic | 16.11 | 18.21 | 18.49 | 17.08 | 18.16 | 15.72 | 13.74 | 11.7 | 10.02 | 9.35 | 9.36 | 19.35 |
| Sargan p value | 0.16 | 0.07 | 0.48 | 0.09 | 0.02 | 0.01 | 0.49 | 0.09 | 0.83 | 0.61 | 0.61 | 0.31 |
| Adjusted $R^2$ | 0.14 | 0.14 | 0.13 | 0.12 | 0.16 | 0.16 | 0.12 | 0.11 | 0.23 | 0.23 | 0.22 | 0.32 |
| *N* | 2,055 | 1,892 | 1,407 | 842 | 1,874 | 1,711 | 1,254 | 751 | 181 | 181 | 153 | 91 |
| | **B. $NO_2$** | | | | | | | | | | | |
| | All | 2km | 1km | .5km | All | 2km | 1km | .5km | All | 2km | 1km | .5km |
| | (1) | (2) | (3) | (4) | (5) | (6) | (7) | (8) | (9) | (10) | (11) | (12) |
| $NO_2$ | $0.41^{**}$ | $0.45^{**}$ | $0.43^{**}$ | $0.48^{*}$ | $0.41^{**}$ | $0.47^{**}$ | $0.66^{**}$ | $0.73^{*}$ | $-0.06$ | $-0.07$ | $-0.10$ | $-0.21$ |
| | (0.18) | (0.19) | (0.22) | (0.27) | (0.19) | (0.23) | (0.31) | (0.38) | (0.13) | (0.13) | (0.13) | (0.15) |
| AME | 1.77 | 1.98 | 1.96 | 2.28 | 1.75 | 2.12 | 3.25 | 3.71 | -0.18 | -0.22 | -0.31 | -0.7 |
| | (0.13, 3.99) | (0.13, 4.4) | (0.08, 5.42) | (-0.11, 7.19) | (0.02, 4) | (-0.07, 5.29) | (0.08, 9.95) | (-0.17, 13.73) | (-0.89, 0.67) | (-0.98, 0.64) | (-1.27, 0.55) | (-1.92, 0.66) |
| Geography | All | All | All | All | Outer-Borough | Outer-Borough | Outer-Borough | Outer-Borough | Manhattan | Manhattan | Manhattan | Manhattan |
| F statistic | 12.76 | 15.25 | 16.12 | 14.47 | 13.29 | 12.95 | 11.1 | 10.16 | 5.32 | 6.75 | 8.4 | 12.32 |
| Sargan p value | 0.16 | 0.06 | 0.43 | 0.08 | 0.02 | 0 | 0.34 | 0.06 | 0.84 | 0.62 | 0.64 | 0.34 |
| Adjusted $R^2$ | 0.13 | 0.13 | 0.12 | 0.10 | 0.16 | 0.16 | 0.11 | 0.10 | 0.23 | 0.24 | 0.22 | 0.33 |
| *N* | 2,055 | 1,892 | 1,407 | 842 | 1,874 | 1,711 | 1,254 | 751 | 181 | 181 | 153 | 91 |
| | **C. NO** | | | | | | | | | | | |
| | All | 2km | 1km | .5km | All | 2km | 1km | .5km | All | 2km | 1km | .5km |
| | (1) | (2) | (3) | (4) | (5) | (6) | (7) | (8) | (9) | (10) | (11) | (12) |
| NO | $0.22^{**}$ | $0.24^{**}$ | $0.25^{**}$ | $0.25^{*}$ | $0.19^{*}$ | 0.22 | $0.36^{**}$ | $0.38^{*}$ | $-0.03$ | $-0.04$ | $-0.06$ | $-0.12$ |
| | (0.11) | (0.11) | (0.13) | (0.15) | (0.11) | (0.14) | (0.19) | (0.21) | (0.07) | (0.07) | (0.07) | (0.09) |
| AME | 1.85 | 2.06 | 2.23 | 2.29 | 1.59 | 1.88 | 3.54 | 3.71 | -0.18 | -0.24 | -0.33 | -0.76 |
| | (-0.19, 4.68) | (0.02, 5.49) | (-0.02, 5.73) | (-0.2, 8.03) | (-0.39, 4.51) | (-0.67, 5.79) | (0.14, 11.49) | (-0.31, 16.38) | (-0.9, 0.61) | (-1.08, 0.59) | (-1.24, 0.7) | (-1.99, 0.65) |
| Geography | All | All | All | All | Outer-Borough | Outer-Borough | Outer-Borough | Outer-Borough | Manhattan | Manhattan | Manhattan | Manhattan |
| F statistic | 9.76 | 11.98 | 12.98 | 12.05 | 10.07 | 9.52 | 7.85 | 7.78 | 6.24 | 7.39 | 8.82 | 12.49 |
| Sargan p value | 0.11 | 0.04 | 0.44 | 0.06 | 0.01 | 0 | 0.28 | 0.05 | 0.84 | 0.63 | 0.64 | 0.36 |
| Adjusted $R^2$ | 0.12 | 0.12 | 0.09 | 0.10 | 0.16 | 0.16 | 0.08 | 0.08 | 0.23 | 0.24 | 0.22 | 0.32 |
| *N* | 2,055 | 1,892 | 1,407 | 842 | 1,874 | 1,711 | 1,254 | 751 | 181 | 181 | 153 | 91 |

*Notes:* The dependent variable in these 2SLS log-linear models is the logged count plus one of deaths measured at the census tract level. The reported coefficients are based on samples that include all tracts within the given Geography and maximum highway distance that include at least 500 people, according to the 2018 5-year ACS, with centroids that lie further than 0.05km from the nearest highway segment. Each model includes the following additional controls: distance to the nearest highway, and indicators for PUMA and nearest weather station. Robust standard errors are reported in parentheses. One, two, and three stars indicate 10 percent, 5 percent, and 1 percent significance, respectively. Average Marginal Effects (AME) are reported with 95% bootstrapped confidence intervals.

### Table S9: Near Distance Analysis on Hospitalizations

**A. $PM_{2.5}$**

| | All | 2km | 1km | .5km | All | 2km | 1km | .5km | All | 2km | 1km | .5km |
|---|---|---|---|---|---|---|---|---|---|---|---|---|
| | (1) | (2) | (3) | (4) | (5) | (6) | (7) | (8) | (9) | (10) | (11) | (12) |
| $PM_{2.5}$ | 0.50 | 0.71* | 0.56 | 0.53 | 0.46 | 0.76* | 1.11* | 0.90 | 0.02 | 0.02 | −0.40 | −0.34 |
| | (0.37) | (0.40) | (0.45) | (0.51) | (0.38) | (0.46) | (0.60) | (0.70) | (0.26) | (0.30) | (0.31) | (0.36) |
| AME | 2.03 | 3.06 | 2.45 | 2.37 | 1.9 | 3.35 | 5.33 | 4.25 | 0.05 | 0.07 | -1.26 | -1.18 |
| | (-1.45, 6.32) | (-0.71, 8.49) | (-1.29, 8.94) | (-1.92, 9.59) | (-1.4, 5.89) | (-0.79, 9.48) | (0.05, 16.43) | (-2.25, 19.37) | (-1.62, 1.97) | (-1.82, 2.26) | (-3.16, 1.63) | (-3.81, 2.51) |
| Geography | All | All | All | All | Outer-Borough | Outer-Borough | Outer-Borough | Outer-Borough | Manhattan | Manhattan | Manhattan | Manhattan |
| F statistic | 16.11 | 18.21 | 18.49 | 17.08 | 18.16 | 15.72 | 13.74 | 11.7 | 10.02 | 9.35 | 9.36 | 19.35 |
| Sargan p value | 0 | 0 | 0.02 | 0.19 | 0 | 0 | 0.13 | 0.31 | 0.52 | 0.46 | 0.19 | 0.11 |
| Adjusted $R^2$ | 0.26 | 0.25 | 0.26 | 0.27 | 0.28 | 0.28 | 0.24 | 0.27 | 0.20 | 0.20 | 0.20 | 0.16 |
| *N* | 2,055 | 1,892 | 1,407 | 842 | 1,874 | 1,711 | 1,254 | 751 | 181 | 181 | 153 | 91 |

**B. $NO_2$**

| | All | 2km | 1km | .5km | All | 2km | 1km | .5km | All | 2km | 1km | .5km |
|---|---|---|---|---|---|---|---|---|---|---|---|---|
| | (1) | (2) | (3) | (4) | (5) | (6) | (7) | (8) | (9) | (10) | (11) | (12) |
| $NO_2$ | 0.27 | 0.36* | 0.27 | 0.25 | 0.23 | 0.36 | 0.50* | 0.43 | 0.04 | 0.03 | −0.17 | −0.17 |
| | (0.18) | (0.19) | (0.21) | (0.25) | (0.18) | (0.22) | (0.30) | (0.35) | (0.13) | (0.15) | (0.14) | (0.17) |
| AME | 2.7 | 3.75 | 2.81 | 2.62 | 2.24 | 3.8 | 5.81 | 4.92 | 0.26 | 0.23 | -1.25 | -1.39 |
| | (-1.04, 8.67) | (-0.13, 10.65) | (-1.43, 10.82) | (-2.48, 11.85) | (-1.85, 6.94) | (-1.01, 10.77) | (-0.58, 18.58) | (-2.44, 20.81) | (-1.76, 2.5) | (-1.88, 3.07) | (-3.47, 1.79) | (-5.33, 2.3) |
| Geography | All | All | All | All | Outer-Borough | Outer-Borough | Outer-Borough | Outer-Borough | Manhattan | Manhattan | Manhattan | Manhattan |
| F statistic | 12.76 | 15.25 | 16.12 | 14.47 | 13.29 | 12.95 | 11.1 | 10.16 | 5.32 | 6.75 | 8.4 | 12.32 |
| Sargan p value | 0 | 0 | 0.02 | 0.17 | 0 | 0 | 0.09 | 0.26 | 0.54 | 0.47 | 0.17 | 0.12 |
| Adjusted $R^2$ | 0.25 | 0.24 | 0.26 | 0.27 | 0.28 | 0.28 | 0.25 | 0.27 | 0.19 | 0.19 | 0.20 | 0.15 |
| *N* | 2,055 | 1,892 | 1,407 | 842 | 1,874 | 1,711 | 1,254 | 751 | 181 | 181 | 153 | 91 |

**C. NO**

| | All | 2km | 1km | .5km | All | 2km | 1km | .5km | All | 2km | 1km | .5km |
|---|---|---|---|---|---|---|---|---|---|---|---|---|
| | (1) | (2) | (3) | (4) | (5) | (6) | (7) | (8) | (9) | (10) | (11) | (12) |
| NO | 0.17 | 0.21* | 0.17 | 0.12 | 0.10 | 0.18 | 0.30* | 0.22 | 0.02 | 0.02 | −0.09 | −0.10 |
| | (0.11) | (0.11) | (0.12) | (0.14) | (0.11) | (0.13) | (0.18) | (0.20) | (0.07) | (0.08) | (0.08) | (0.09) |
| AME | 3.18 | 4.26 | 3.52 | 2.56 | 1.87 | 3.61 | 6.79 | 4.93 | 0.22 | 0.25 | -1.3 | -1.56 |
| | (-1.46, 9.76) | (-0.72, 13.36) | (-1.3, 13.86) | (-2.7, 13.49) | (-2.75, 7.94) | (-2.37, 13.19) | (-0.89, 25.02) | (-3.02, 22.23) | (-1.75, 2.28) | (-1.96, 3.41) | (-3.73, 1.99) | (-5.77, 3.15) |
| Geography | All | All | All | All | Outer-Borough | Outer-Borough | Outer-Borough | Outer-Borough | Manhattan | Manhattan | Manhattan | Manhattan |
| F statistic | 9.76 | 11.98 | 12.98 | 12.05 | 10.07 | 9.52 | 7.85 | 7.78 | 6.24 | 7.39 | 8.82 | 12.49 |
| Sargan p value | 0 | 0 | 0.02 | 0.15 | 0 | 0 | 0.1 | 0.23 | 0.53 | 0.47 | 0.17 | 0.13 |
| Adjusted $R^2$ | 0.24 | 0.22 | 0.23 | 0.27 | 0.28 | 0.27 | 0.21 | 0.26 | 0.19 | 0.19 | 0.19 | 0.15 |
| *N* | 2,055 | 1,892 | 1,407 | 842 | 1,874 | 1,711 | 1,254 | 751 | 181 | 181 | 153 | 91 |

*Notes:* The dependent variable in these 2SLS log-linear models is the logged count plus one of hospitalizations measured at the census tract level. The reported coefficients are based on samples that include all tracts within the given Geography and maximum highway distance that include at least 500 people, according to the 2018 5-year ACS, with centroids that lie further than 0.05km from the nearest highway segment. Each model includes the following additional controls: distance to the nearest highway, and indicators for PUMA and nearest weather station. Robust standard errors are reported in parentheses. One, two, and three stars indicate 10 percent, 5 percent, and 1 percent significance, respectively. Average Marginal Effects (AME) are reported with 95% bootstrapped confidence intervals.

#### 3.4 Race and Age Stratified Regression

We also explore whether the effects of long-term air pollution concentration on the intensity of COVID-19 disease differ based on demographics. To do so, we stratify our samples and estimate equation (2) for each race and age group separately. The outcome variables are log transformed counts of deaths and hospitalization for each age and race in each census tract. The race groups include White, Hispanic/Latino, Black/African American, and Asian/Pacific Islander. The age groups include 18-65, 65-74, and 75+. In these regressions, we restrict observations to tracts with a population greater than 200 in that age or race stratum, as tracts with few individuals in a given stratum could bias results. In general, we encourage careful interpretation of these results, as populations vary widely between tracts. Table S10 reports the result of the race stratified regressions and Table S11 reports the result of the age stratified regressions.

### Table S10: Race Stratified Regressions

**A. Deaths**

| | (1) | (2) | (3) | (4) | (5) | (6) | (7) | (8) | (9) | (10) | (11) | (12) |
|---|---|---|---|---|---|---|---|---|---|---|---|---|
| | $PM_{2.5}$ | $PM_{2.5}$ | $PM_{2.5}$ | $PM_{2.5}$ | $NO_2$ | $NO_2$ | $NO_2$ | $NO_2$ | NO | NO | NO | NO |
| AQ | 1.42*** | 0.84 | −0.71 | 0.38 | 0.68*** | 0.38 | −0.27 | 0.20 | 0.39*** | 0.19 | −0.03 | 0.11 |
| | (0.52) | (0.55) | (1.09) | (0.37) | (0.26) | (0.27) | (0.47) | (0.20) | (0.15) | (0.15) | (0.22) | (0.11) |
| Race/Ethnicity | White | Hispanic/Latino | Black/African American | Asian/Pacific Islander | White | Hispanic/Latino | Black/African American | Asian/Pacific Islander | White | Hispanic/Latino | Black/African American | Asian/Pacific Islander |
| Adjusted $R^2$ | 0.08 | 0.40 | 0.24 | 0.19 | 0.05 | 0.41 | 0.24 | 0.20 | 0.02 | 0.40 | 0.25 | 0.19 |
| $N$ | 858 | 1,111 | 684 | 711 | 858 | 1,111 | 684 | 711 | 858 | 1,111 | 684 | 711 |

**B. Hospitalizations**

| | (1) | (2) | (3) | (4) | (5) | (6) | (7) | (8) | (9) | (10) | (11) | (12) |
|---|---|---|---|---|---|---|---|---|---|---|---|---|
| | $PM_{2.5}$ | $PM_{2.5}$ | $PM_{2.5}$ | $PM_{2.5}$ | $NO_2$ | $NO_2$ | $NO_2$ | $NO_2$ | NO | NO | NO | NO |
| AQ | 1.40*** | 0.88 | 0.08 | 0.49 | 0.64** | 0.40 | 0.06 | 0.27 | 0.37** | 0.21 | 0.17 | 0.14 |
| | (0.54) | (0.58) | (1.18) | (0.44) | (0.27) | (0.28) | (0.52) | (0.23) | (0.16) | (0.16) | (0.26) | (0.13) |
| Race/Ethnicity | White | Hispanic/Latino | Black/African American | Asian/Pacific Islander | White | Hispanic/Latino | Black/African American | Asian/Pacific Islander | White | Hispanic/Latino | Black/African American | Asian/Pacific Islander |
| Adjusted $R^2$ | 0.09 | 0.47 | 0.31 | 0.30 | 0.08 | 0.47 | 0.31 | 0.31 | 0.06 | 0.47 | 0.30 | 0.30 |
| $N$ | 858 | 1,111 | 684 | 711 | 858 | 1,111 | 684 | 711 | 858 | 1,111 | 684 | 711 |

*Notes:* The dependent variable in these 2SLS log-linear models is the logged count plus one of our COVID-19 outcome of interest (deaths or hospitalizations) measured at the census tract level. The reported coefficients are based on samples that include all tracts within the Outer-Boroughs sample that include at least 200 people of the modeled race/ethnicity, according to the 2018 5-year ACS, with centroids that lie between 0.05 and 1km from the nearest highway segment. Each model includes the following additional controls: distance to the nearest highway, and indicators for PUMA and nearest weather station. Robust standard errors are reported in parentheses. One, two, and three stars indicate 10 percent, 5 percent, and 1 percent significance, respectively.

Table S11: Age Stratified Regressions

**A. Deaths**

| | (1) | (2) | (3) | (4) | (5) | (6) | (7) | (8) | (9) |
|---|---|---|---|---|---|---|---|---|---|
| | $PM_{2.5}$ | $PM_{2.5}$ | $PM_{2.5}$ | $NO_2$ | $NO_2$ | $NO_2$ | NO | NO | NO |
| AQ | 0.68 | −0.20 | 1.36*** | 0.35 | −0.14 | 0.72*** | 0.21 | −0.13 | 0.39*** |
| | (0.56) | (0.72) | (0.49) | (0.28) | (0.35) | (0.27) | (0.17) | (0.17) | (0.14) |
| Age | 18-64 | 65-74 | 75+ | 18-64 | 65-74 | 75+ | 18-64 | 65-74 | 75+ |
| Adjusted $R^2$ | 0.24 | 0.20 | 0.03 | 0.24 | 0.20 | 0.001 | 0.22 | 0.20 | −0.01 |
| *N* | 1,255 | 635 | 711 | 1,255 | 635 | 711 | 1,255 | 635 | 711 |

**B. Hospitalizations**

| | (1) | (2) | (3) | (4) | (5) | (6) | (7) | (8) | (9) |
|---|---|---|---|---|---|---|---|---|---|
| | $PM_{2.5}$ | $PM_{2.5}$ | $PM_{2.5}$ | $NO_2$ | $NO_2$ | $NO_2$ | NO | NO | NO |
| AQ | 1.33** | −0.13 | 0.88* | 0.63* | −0.04 | 0.46* | 0.39* | −0.02 | 0.25* |
| | (0.65) | (0.68) | (0.48) | (0.33) | (0.34) | (0.26) | (0.20) | (0.17) | (0.14) |
| Age | 18-64 | 65-74 | 75+ | 18-64 | 65-74 | 75+ | 18-64 | 65-74 | 75+ |
| Adjusted $R^2$ | 0.26 | 0.26 | 0.13 | 0.26 | 0.26 | 0.12 | 0.20 | 0.26 | 0.12 |
| *N* | 1,255 | 635 | 711 | 1,255 | 635 | 711 | 1,255 | 635 | 711 |

*Notes:* The dependent variable in these 2SLS log-linear models is the logged count plus one of our COVID-19 outcome of interest (deaths or hospitalizations) measured at the census tract level. The reported coefficients are based on samples that include all tracts within the Outer Borough sample that include at least 200 people of the modeled age group, according to the 2018 5-year ACS, with centroids that lie between 0.05 and 1km from the nearest highway segment. Each model includes the following additional controls: distance to the nearest highway, and indicators for PUMA and nearest weather station. Robust standard errors are reported in parentheses. One, two, and three stars indicate 10 percent, 5 percent, and 1 percent significance, respectively.

#### 3.5 Count Based vs Rate Based Measures

Most previous studies of the relationship between air quality and covid outcomes have normalized the covid outcomes by population. However given the discussion surrounding Figure S3, we show that tract population is an inappropriate denominator given the number of people who left the city in some tracts. To further explore the shortcomings of rate based measures in this context, we show our results using rate based dependent variables in Table S12. Table S12, panel A shows the results using our instrumental variable approach on a dependent variable of log(Deaths*100,000/Tract Population + 1). We also use tract population weights in these specifications.

Our estimates are still high, showing approximately a 22% increase in deaths, however they are not statistically significant. In Table S12, panel B we adjust the population by the fraction of Safegraph devices that left the city between weeks 10 and 20, however this mostly affects the results in the Manhattan sample. This adjustment is likely unsatisfactory because older individuals are less likely to be represented in the Safegraph data and more likely to be susceptible to COVID-19 and poor air quality.

Table S12 panel C and D show the results of regressions on log mortality rates and log hospitalization rates with a rich set of control variables, but using an OLS approach that does not account for omitted variable bias. These tables show a negative relationship between air quality and COVID-19 outcomes.

Table S12: Rate Based Measures

| | | | | | | | | | |
|---|---|---|---|---|---|---|---|---|---|
| | **A. Log mortality rate - IV- unadjusted** | | | | | | | | |
| | (1) | (2) | (3) | (4) | (5) | (6) | (7) | (8) | (9) |
| | $PM_{2.5}$ | $PM_{2.5}$ | $PM_{2.5}$ | $NO_2$ | $NO_2$ | $NO_2$ | NO | NO | NO |
| AQ | 0.22 | 0.15 | 0.04 | 0.09 | 0.09 | −0.001 | 0.03 | 0.02 | −0.002 |
| | (0.39) | (0.57) | (0.42) | (0.18) | (0.28) | (0.18) | (0.09) | (0.14) | (0.10) |
| Geography | Citywide | Outer Boroughs | Manhattan | Citywide | Outer Boroughs | Manhattan | Citywide | Outer Boroughs | Manhattan |
| Adjusted $R^2$ | 0.27 | 0.21 | 0.16 | 0.28 | 0.21 | 0.16 | 0.28 | 0.21 | 0.16 |
| $N$ | 1,407 | 1,254 | 153 | 1,407 | 1,254 | 153 | 1,407 | 1,254 | 153 |
| | **B. Log mortality rate -IV- adjusted** | | | | | | | | |
| | (1) | (2) | (3) | (4) | (5) | (6) | (7) | (8) | (9) |
| | $PM_{2.5}$ | $PM_{2.5}$ | $PM_{2.5}$ | $NO_2$ | $NO_2$ | $NO_2$ | NO | NO | NO |
| AQ | 0.22 | 0.15 | 0.10 | 0.07 | 0.04 | 0.003 | 0.01 | −0.01 | −0.001 |
| | (0.43) | (0.58) | (0.44) | (0.20) | (0.29) | (0.19) | (0.11) | (0.15) | (0.10) |
| Geography | Citywide | Outer Boroughs | Manhattan | Citywide | Outer Boroughs | Manhattan | Citywide | Outer Boroughs | Manhattan |
| Adjusted $R^2$ | 0.20 | 0.19 | 0.07 | 0.20 | 0.19 | 0.08 | 0.20 | 0.19 | 0.08 |
| $N$ | 1,405 | 1,252 | 153 | 1,405 | 1,252 | 153 | 1,405 | 1,252 | 153 |

### Table S12: Rate Based Measures (continued)

| | C. Log mortality rate - OLS w/ demographic controls | | | | | | | | |
|---|---|---|---|---|---|---|---|---|---|
| | (1) | (2) | (3) | (4) | (5) | (6) | (7) | (8) | (9) |
| | $PM_{2.5}$ | $PM_{2.5}$ | $PM_{2.5}$ | $NO_2$ | $NO_2$ | $NO_2$ | NO | NO | NO |
| AQ | 9.17 | 22.33 | −0.93 | 6.38 | 13.73 | 1.04 | 3.26 | 7.15 | −0.03 |
| | (15.49) | (24.38) | (13.83) | (6.47) | (11.19) | (5.64) | (3.33) | (5.67) | (2.95) |
| Population | −0.01*** | −0.01** | −0.01** | −0.01*** | −0.01** | −0.01** | −0.01*** | −0.01** | −0.01** |
| | (0.003) | (0.003) | (0.003) | (0.003) | (0.003) | (0.003) | (0.003) | (0.003) | (0.003) |
| White | 165.91*** | 151.32** | 278.08** | 166.65*** | 151.97** | 281.28** | 166.48*** | 151.52** | 278.80** |
| | (57.01) | (59.36) | (122.35) | (57.05) | (59.21) | (123.53) | (57.07) | (59.09) | (122.72) |
| Black/African American | 199.18*** | 177.73*** | 681.31* | 198.72*** | 176.94*** | 681.42* | 198.61*** | 176.56*** | 681.33* |
| | (44.53) | (44.68) | (373.22) | (44.26) | (44.16) | (372.85) | (44.13) | (43.80) | (373.24) |
| Hispanic/Latino | 129.08*** | 108.51** | 176.18 | 129.24*** | 108.72** | 179.43 | 129.00*** | 107.86** | 176.97 |
| | (39.13) | (43.06) | (121.54) | (39.33) | (43.33) | (121.59) | (39.34) | (43.56) | (121.73) |
| Age 65-74 | −1,598.28*** | −1,680.40*** | −630.43 | −1,597.77*** | −1,683.96*** | −615.46 | −1,598.71*** | −1,685.13*** | −626.92 |
| | (418.20) | (456.33) | (419.92) | (417.68) | (457.55) | (419.97) | (417.64) | (458.35) | (422.16) |
| Age 75+ | −436.83** | −449.33** | −366.17 | −434.21** | −444.29** | −364.55 | −435.16** | −445.90** | −366.00 |
| | (184.65) | (202.45) | (241.28) | (184.43) | (202.30) | (240.75) | (184.52) | (202.15) | (241.12) |
| High School Education | 309.21*** | 276.50*** | 647.13*** | 307.75*** | 276.08*** | 643.67*** | 308.06*** | 276.41*** | 646.39*** |
| | (101.30) | (97.77) | (242.50) | (101.06) | (97.82) | (241.92) | (100.96) | (97.94) | (242.72) |
| Less than HS education | 342.08*** | 322.06*** | 452.80*** | 340.17*** | 320.53*** | 454.75*** | 340.85*** | 322.24*** | 453.24*** |
| | (86.94) | (94.24) | (162.91) | (86.59) | (94.03) | (163.43) | (86.52) | (94.26) | (162.75) |
| Income per capita | −0.0003 | −0.0004 | −0.0002 | −0.0003 | −0.0004 | −0.0003 | −0.0003 | −0.0004 | −0.0002 |
| | (0.0003) | (0.001) | (0.0004) | (0.0003) | (0.001) | (0.0004) | (0.0003) | (0.001) | (0.0004) |
| Owner Occupied | −40.51 | −62.96* | 118.38** | −39.98 | −62.23 | 118.87** | −40.67 | −63.67* | 118.52** |
| | (35.37) | (38.16) | (48.86) | (35.13) | (37.93) | (49.10) | (35.01) | (37.80) | (48.99) |
| Receiving Public Assistance | −186.85 | −213.53 | −701.52 | −187.79 | −215.44 | −699.05 | −185.29 | −210.49 | −700.76 |
| | (210.55) | (229.27) | (484.23) | (210.05) | (229.12) | (483.58) | (209.86) | (228.36) | (484.39) |
| Geography | Citywide | Outer Boroughs | Manhattan | Citywide | Outer Boroughs | Manhattan | Citywide | Outer Boroughs | Manhattan |
| Adjusted $R^2$ | 0.25 | 0.20 | 0.47 | 0.25 | 0.20 | 0.47 | 0.25 | 0.20 | 0.47 |
| *N* | 1,404 | 1,251 | 153 | 1,404 | 1,251 | 153 | 1,404 | 1,251 | 153 |

Table S12: Rate Based Measures (continued)

| | D. Log hospitalization rate - OLS w/ demographic controls | | | | | | | | |
|---|---|---|---|---|---|---|---|---|---|
| | (1) | (2) | (3) | (4) | (5) | (6) | (7) | (8) | (9) |
| | $PM_{2.5}$ | $PM_{2.5}$ | $PM_{2.5}$ | $NO_2$ | $NO_2$ | $NO_2$ | NO | NO | NO |
| AQ | −6.64 | −32.81 | 29.59 | 5.15 | −3.39 | 14.78* | −1.01 | −8.08 | 7.51* |
| | (26.37) | (44.29) | (20.31) | (11.65) | (20.78) | (8.33) | (6.26) | (10.78) | (4.35) |
| Population | −0.01*** | −0.02*** | −0.01* | −0.01*** | −0.02*** | −0.01* | −0.01*** | −0.02*** | −0.01* |
| | (0.005) | (0.01) | (0.004) | (0.005) | (0.01) | (0.004) | (0.005) | (0.01) | (0.004) |
| White | 332.02*** | 319.18*** | 481.04*** | 332.54*** | 316.17*** | 486.89*** | 331.83*** | 318.17*** | 484.76*** |
| | (83.72) | (86.75) | (161.69) | (83.72) | (86.79) | (162.59) | (83.67) | (86.78) | (162.32) |
| Black/African American | 440.14*** | 410.51*** | 1,205.93** | 437.93*** | 406.26*** | 1,206.25** | 439.81*** | 410.60*** | 1,207.11** |
| | (71.83) | (74.69) | (507.58) | (71.82) | (74.49) | (505.79) | (71.63) | (74.20) | (506.91) |
| Hispanic/Latino | 493.65*** | 475.88*** | 667.79*** | 492.30*** | 470.29*** | 670.69*** | 493.27*** | 475.08*** | 671.20*** |
| | (74.43) | (83.66) | (191.85) | (74.75) | (83.72) | (192.34) | (74.56) | (83.40) | (192.89) |
| Age 65-74 | −608.56 | −640.57 | 15.77 | −605.14 | −648.47 | 36.13 | −607.61 | −637.66 | 28.80 |
| | (426.09) | (461.68) | (586.88) | (425.78) | (462.50) | (583.44) | (425.87) | (462.13) | (587.22) |
| Age 75+ | −169.49 | −306.29 | 428.66 | −167.66 | −309.87 | 441.18 | −170.08 | −310.81 | 434.41 |
| | (408.54) | (458.73) | (634.47) | (408.06) | (457.89) | (635.89) | (408.22) | (458.61) | (633.81) |
| High School Education | 475.09*** | 385.90** | 106.39 | 473.33*** | 388.40** | 99.32 | 475.29*** | 386.66** | 97.45 |
| | (140.28) | (155.89) | (388.25) | (139.99) | (155.90) | (386.78) | (139.91) | (156.05) | (387.84) |
| Less than HS education | 668.48*** | 561.03*** | 815.85*** | 664.64*** | 561.81*** | 819.85*** | 668.23*** | 560.93*** | 816.65*** |
| | (191.16) | (211.28) | (259.78) | (191.89) | (211.76) | (260.15) | (191.65) | (211.14) | (260.08) |
| Income per capita | −0.001*** | −0.003*** | −0.001** | −0.001*** | −0.003*** | −0.001** | −0.001*** | −0.003*** | −0.001** |
| | (0.0005) | (0.001) | (0.0004) | (0.0005) | (0.001) | (0.0004) | (0.0005) | (0.001) | (0.0004) |
| Owner Occupied | −188.24*** | −217.22*** | 120.53* | −186.08** | −214.79*** | 119.61* | −187.72*** | −215.69*** | 120.27* |
| | (72.25) | (79.79) | (68.32) | (72.35) | (80.12) | (68.01) | (72.06) | (79.66) | (67.96) |
| Receiving Public Assistance | −9.47 | −74.35 | −329.60 | −7.72 | −71.53 | −335.49 | −9.27 | −77.14 | −329.40 |
| | (275.93) | (295.70) | (739.53) | (275.85) | (295.86) | (738.24) | (275.77) | (295.62) | (738.88) |
| Geography | Citywide | Outer Boroughs | Manhattan | Citywide | Outer Boroughs | Manhattan | Citywide | Outer Boroughs | Manhattan |
| Adjusted $R^2$ | 0.50 | 0.45 | 0.61 | 0.50 | 0.45 | 0.61 | 0.50 | 0.45 | 0.61 |
| *N* | 1,404 | 1,251 | 153 | 1,404 | 1,251 | 153 | 1,404 | 1,251 | 153 |

The difference in effects found resulting from choice of model and dependent variable can be visualized in Figure S6. OLS models that do not account for the endogeneity of air pollution or the measurement error in dependent variables find coefficients close to zero, even when controls are included and population size is adjusted for departures from the city using Safegraph data. 2SLS models account for endogeneity, however if rate variables are used, there is a big difference in the effect size depending on whether or not control variables are included. This shows how the measurement error in the dependent variable that is correlated with the dependent variable can still bias instrumental variable estimates. In our regressions on the count variables, there is a much smaller difference between the specifications with and without control variables, and a much larger effect in general. Our preferred specifications do not include control variables, which can bias estimation by conditioning on post-treatment variables.

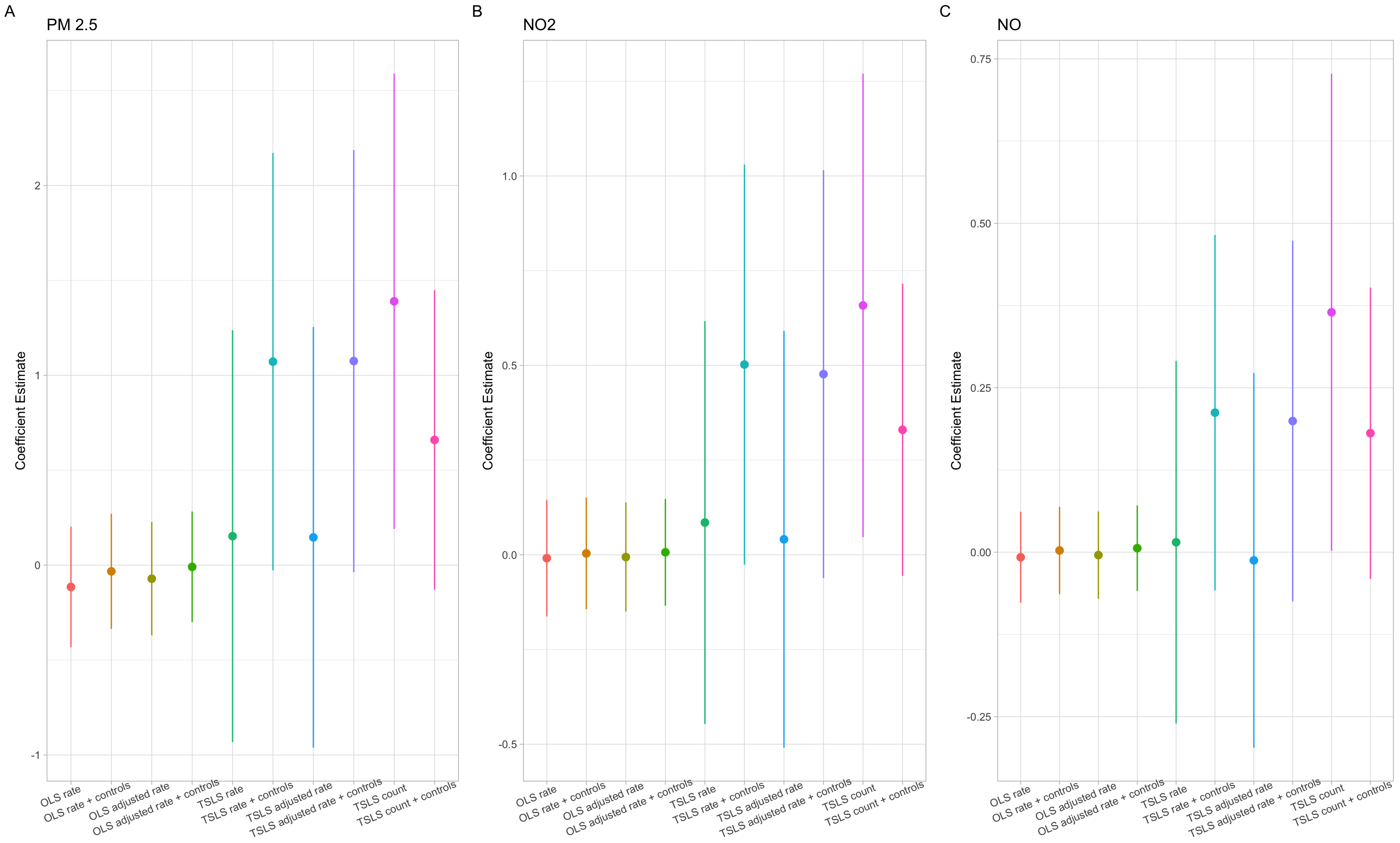

Figure S6: Choice of Model and Dependent Variable

##### 3.6 Quantile Regression

We also perform quantile regressions to explore how the effects of air quality operate across the distribution of tracts by COVID-19 deaths. Figure S6 shows the quantile regression results with the same set of tract restrictions used in our main specification. We find that the effect of air quality is most pronounced in the tracts with the highest number of deaths. This is likely because high populations and high infection rates make it easier to identify the noisy signal from air quality.

The 90th percentile tract in our Outer Borough sample has 20 deaths. These tracts have a higher proportion of Black/African American residents (.24 vs .21), Hispanic/Latino residents (.49 vs .31), and lower per capita income ($21,782 vs $29,373) when compared with the rest of the Outer Borough sample.

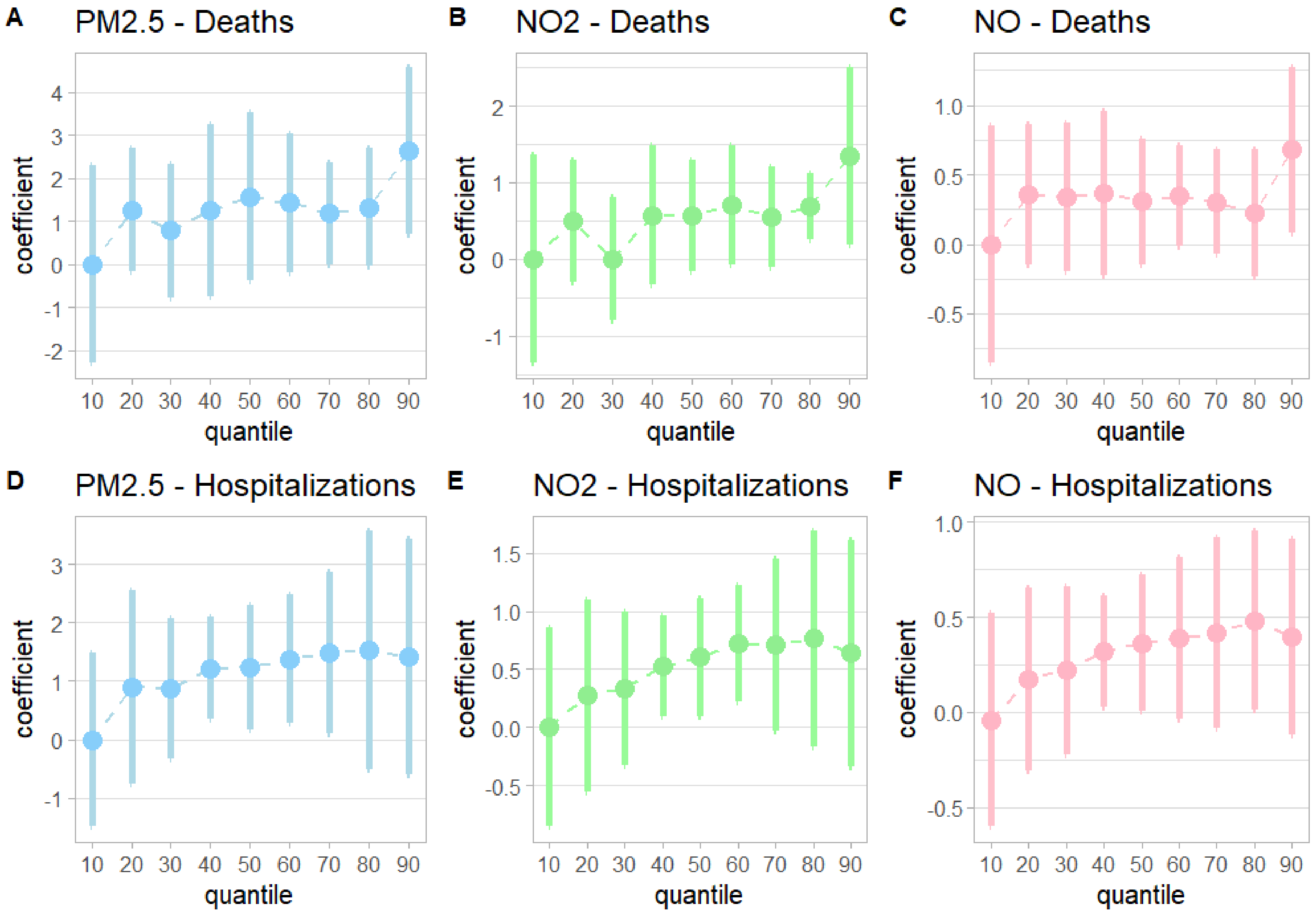

Figure S7: Quantile Regression Analysis

#### 3.7 Demographic Controls

In addition to the robustness checks previously mentioned, we also run versions of equations (1) and (2) with demographic control variables from ACS and Safegraph. While this runs the risk of biasing estimation by conditioning on post-treatment variables, as air quality has been shown to affect educational achievement and income, for instance (Zhang et al. 2010), all of our estimates remain positive, large, and statistically significant in the citywide regressions (Table S13, Panel A and B).

Panel A shows results from a simple set of control variables including population, race, age, education, and a few economic variables, which are comparable to the sets of controls used in other observational studies. Panel B adds additional controls which may be more important in the NYC setting, including Institutionalized - for the fraction of the population living in group quarters, median tenure, occupations, public transport, and the change in observed cell devices during the first wave of the pandemic.

### Table S13: IV with Demographics Controls

| | A. Deaths - demographic controls | | | | | | | | |
|---|---|---|---|---|---|---|---|---|---|
| | (1) | (2) | (3) | (4) | (5) | (6) | (7) | (8) | (9) |
| | $PM_{2.5}$ | $PM_{2.5}$ | $PM_{2.5}$ | $NO_2$ | $NO_2$ | $NO_2$ | NO | NO | NO |
| AQ | 0.66** | 0.66 | 0.14 | 0.32** | 0.33* | 0.03 | 0.17** | 0.18 | 0.01 |
| | (0.30) | (0.40) | (0.23) | (0.14) | (0.20) | (0.10) | (0.08) | (0.11) | (0.06) |
| Tract Population | 0.0002*** | 0.0002*** | 0.0001*** | 0.0002*** | 0.0002*** | 0.0001*** | 0.0002*** | 0.0002*** | 0.0001*** |
| | (0.0000) | (0.0000) | (0.0000) | (0.0000) | (0.0000) | (0.0000) | (0.0000) | (0.0000) | (0.0000) |
| % White | 0.36** | 0.25 | 1.81*** | 0.40*** | 0.28* | 1.73*** | 0.39** | 0.27* | 1.72*** |
| | (0.15) | (0.16) | (0.63) | (0.15) | (0.15) | (0.63) | (0.15) | (0.16) | (0.62) |
| % Black/African American | 0.59*** | 0.44*** | 1.80** | 0.59*** | 0.44*** | 1.80** | 0.58*** | 0.43*** | 1.80** |
| | (0.14) | (0.15) | (0.75) | (0.14) | (0.15) | (0.75) | (0.14) | (0.15) | (0.75) |
| % Hispanic/Latino | 0.49*** | 0.31* | 1.70** | 0.52*** | 0.34** | 1.62** | 0.50*** | 0.32* | 1.61** |
| | (0.14) | (0.17) | (0.75) | (0.14) | (0.16) | (0.74) | (0.14) | (0.17) | (0.75) |
| % Age 65-74 | −4.16*** | −4.86*** | 0.72 | −4.18*** | −4.90*** | 0.35 | −4.22*** | −4.95*** | 0.30 |
| | (0.80) | (0.83) | (2.63) | (0.80) | (0.84) | (2.64) | (0.81) | (0.86) | (2.65) |
| % Age 75+ | −2.14*** | −2.40*** | −0.80 | −2.01*** | −2.27*** | −0.80 | −2.05*** | −2.30*** | −0.82 |
| | (0.60) | (0.63) | (1.21) | (0.61) | (0.63) | (1.21) | (0.61) | (0.63) | (1.19) |
| % Education: less than high school | 0.90*** | 0.55** | 2.20* | 0.84*** | 0.53* | 2.27* | 0.85*** | 0.54* | 2.28* |
| | (0.27) | (0.28) | (1.31) | (0.27) | (0.28) | (1.30) | (0.27) | (0.28) | (1.30) |
| % Education: High School | 0.99*** | 0.70*** | 3.22*** | 0.93*** | 0.66** | 3.17*** | 0.95*** | 0.70** | 3.16*** |
| | (0.24) | (0.27) | (0.83) | (0.25) | (0.27) | (0.82) | (0.25) | (0.27) | (0.82) |
| Income Per Capita | −0.0000** | −0.0000*** | −0.0000 | −0.0000** | −0.0000*** | −0.0000 | −0.0000** | −0.0000*** | −0.0000 |
| | (0.0000) | (0.0000) | (0.0000) | (0.0000) | (0.0000) | (0.0000) | (0.0000) | (0.0000) | (0.0000) |
| Owner Occupied | −0.03 | −0.15 | 0.83*** | −0.03 | −0.14 | 0.81*** | −0.06 | −0.18 | 0.81*** |
| | (0.12) | (0.13) | (0.31) | (0.12) | (0.13) | (0.31) | (0.12) | (0.12) | (0.31) |
| Received Public Assistance | 0.26 | −0.19 | −0.41 | 0.17 | −0.24 | −0.51 | 0.31 | −0.12 | −0.51 |
| | (0.47) | (0.49) | (2.44) | (0.47) | (0.50) | (2.44) | (0.48) | (0.50) | (2.46) |
| Geography | Citywide | Outer Boroughs | Manhattan | Citywide | Outer Boroughs | Manhattan | Citywide | Outer Boroughs | Manhattan |
| Adjusted $R^2$ | 0.52 | 0.54 | 0.57 | 0.52 | 0.54 | 0.58 | 0.51 | 0.53 | 0.58 |
| *N* | 1,404 | 1,251 | 153 | 1,404 | 1,251 | 153 | 1,404 | 1,251 | 153 |

*Notes:*

***Significant at the 1 percent level.
**Significant at the 5 percent level.
*Significant at the 10 percent level.

### Table S13: IV with Demographics Controls (continued)

| | B. Deaths - additional demographic controls | | | | | | | | |
|---|---|---|---|---|---|---|---|---|---|
| | (1) | (2) | (3) | (4) | (5) | (6) | (7) | (8) | (9) |
| | $PM_{2.5}$ | $PM_{2.5}$ | $PM_{2.5}$ | $NO_2$ | $NO_2$ | $NO_2$ | NO | NO | NO |
| AQ | 0.62** | 0.53 | 0.13 | 0.30** | 0.27 | 0.003 | 0.16** | 0.15 | −0.002 |
| | (0.30) | (0.40) | (0.28) | (0.14) | (0.20) | (0.13) | (0.08) | (0.11) | (0.07) |
| Tract Population | 0.0002*** | 0.0002*** | 0.0001*** | 0.0002*** | 0.0002*** | 0.0001*** | 0.0002*** | 0.0002*** | 0.0001*** |
| | (0.0000) | (0.0000) | (0.0000) | (0.0000) | (0.0000) | (0.0000) | (0.0000) | (0.0000) | (0.0000) |
| % White | 0.25 | 0.16 | 1.46** | 0.30* | 0.19 | 1.41** | 0.28* | 0.17 | 1.40** |
| | (0.16) | (0.17) | (0.64) | (0.16) | (0.16) | (0.63) | (0.16) | (0.17) | (0.63) |
| % Black/African American | 0.47*** | 0.39*** | 1.64** | 0.48*** | 0.39*** | 1.62* | 0.47*** | 0.38*** | 1.62* |
| | (0.14) | (0.14) | (0.82) | (0.14) | (0.14) | (0.84) | (0.14) | (0.14) | (0.84) |
| % Hispanic/Latino | 0.48*** | 0.36** | 1.59** | 0.50*** | 0.39** | 1.49** | 0.48*** | 0.37** | 1.48** |
| | (0.15) | (0.16) | (0.75) | (0.15) | (0.16) | (0.74) | (0.15) | (0.16) | (0.74) |
| % Age 65-74 | −3.80*** | −4.33*** | 0.56 | −3.83*** | −4.38*** | 0.18 | −3.87*** | −4.41*** | 0.14 |
| | (0.79) | (0.84) | (2.72) | (0.79) | (0.85) | (2.71) | (0.80) | (0.86) | (2.73) |
| % Age 75+ | −2.44*** | −2.73*** | 0.47 | −2.29*** | −2.63*** | 0.38 | −2.34*** | −2.67*** | 0.36 |
| | (0.62) | (0.65) | (1.65) | (0.63) | (0.66) | (1.66) | (0.63) | (0.65) | (1.65) |
| % Education: less than high school | 0.81*** | 0.47* | 2.03 | 0.76*** | 0.45 | 2.38 | 0.77*** | 0.46* | 2.42 |
| | (0.27) | (0.27) | (1.78) | (0.27) | (0.28) | (1.80) | (0.27) | (0.28) | (1.81) |
| % Education: High School | 0.70*** | 0.41 | 2.67** | 0.66** | 0.37 | 2.67** | 0.68** | 0.41 | 2.66** |
| | (0.27) | (0.28) | (1.05) | (0.27) | (0.28) | (1.05) | (0.27) | (0.28) | (1.04) |
| Income Per Capita | −0.0000 | −0.0000*** | −0.0000* | −0.0000 | −0.0000*** | −0.0000 | −0.0000 | −0.0000*** | −0.0000 |
| | (0.0000) | (0.0000) | (0.0000) | (0.0000) | (0.0000) | (0.0000) | (0.0000) | (0.0000) | (0.0000) |
| Owner Occupied | −0.27* | −0.39*** | 0.77** | −0.26* | −0.39*** | 0.78** | −0.29** | −0.42*** | 0.78** |
| | (0.14) | (0.14) | (0.35) | (0.14) | (0.14) | (0.35) | (0.14) | (0.14) | (0.34) |
| Received Public Assistance | 0.06 | −0.28 | −0.87 | −0.04 | −0.34 | −0.83 | 0.11 | −0.22 | −0.82 |
| | (0.47) | (0.49) | (2.52) | (0.47) | (0.49) | (2.54) | (0.48) | (0.50) | (2.54) |
| Median Tenure | 0.02*** | 0.02*** | −0.005 | 0.02*** | 0.02*** | −0.01 | 0.02*** | 0.02*** | −0.01 |
| | (0.01) | (0.01) | (0.02) | (0.01) | (0.01) | (0.02) | (0.01) | (0.01) | (0.02) |
| Institutionalized | 0.84*** | 0.93*** | −0.96 | 0.81*** | 0.91*** | −1.02 | 0.83*** | 0.93*** | −1.02 |
| | (0.29) | (0.31) | (0.84) | (0.29) | (0.31) | (0.84) | (0.29) | (0.31) | (0.83) |
| Service Occupation | −0.28 | −0.61** | 0.96 | −0.28 | −0.59** | 0.89 | −0.32 | −0.63** | 0.89 |
| | (0.28) | (0.29) | (1.18) | (0.28) | (0.28) | (1.16) | (0.29) | (0.29) | (1.16) |
| Uninsured | 0.22 | 0.44 | −0.59 | 0.21 | 0.43 | −0.56 | 0.26 | 0.47 | −0.56 |
| | (0.38) | (0.39) | (2.00) | (0.38) | (0.39) | (1.99) | (0.39) | (0.40) | (1.98) |
| Public Transport | 0.40** | 0.34* | −0.23 | 0.43** | 0.34* | −0.42 | 0.42** | 0.32 | −0.44 |
| | (0.18) | (0.19) | (0.62) | (0.19) | (0.19) | (0.68) | (0.19) | (0.19) | (0.66) |
| Change in cell devices - week 10 to week 20 | 0.29** | 0.29** | −0.33 | 0.28** | 0.29** | −0.35 | 0.27** | 0.29** | −0.35 |
| | (0.12) | (0.12) | (0.33) | (0.12) | (0.12) | (0.32) | (0.12) | (0.12) | (0.32) |
| Geography | Citywide | Outer Boroughs | Manhattan | Citywide | Outer Boroughs | Manhattan | Citywide | Outer Boroughs | Manhattan |
| Adjusted $R^2$ | 0.54 | 0.56 | 0.55 | 0.53 | 0.56 | 0.56 | 0.53 | 0.56 | 0.56 |
| *N* | 1,394 | 1,246 | 148 | 1,394 | 1,246 | 148 | 1,394 | 1,246 | 148 |

*Notes:*

***Significant at the 1 percent level.
**Significant at the 5 percent level.
*Significant at the 10 percent level.

##### 3.8 Additional Robustness Checks

Table S14 reports additional robustness checks. Panel A reports the estimates from equation (2) using 100 rather than 500 as an alternative cutoff for tract population. Panel B reports the estimates from equation (2) using tracts between 0.2 and 2km rather than 0.05 and 1km as the cutoff for distance to the nearest highway. Panel C reports the estimates from equation (2) using pollution interpolated from the closest monitor rather than inverse distance weighted averages. Panel D reports the estimates trimming the top 5% and bottom 5% of tracts by population. Panel E reports results excluding wind speed as an instrument, only using the downwind measures.

Table S14: Additional Robustness Checks

**A. Deaths - Population cutoff 100**

| | (1) | (2) | (3) | (4) | (5) | (6) | (7) | (8) | (9) |
|---|---|---|---|---|---|---|---|---|---|
| | $PM_{2.5}$ | $PM_{2.5}$ | $PM_{2.5}$ | $NO_2$ | $NO_2$ | $NO_2$ | NO | NO | NO |
| AQ | 1.05** | 1.47** | 0.07 | 0.50** | 0.70** | 0.03 | 0.29** | 0.39** | 0.01 |
| | (0.47) | (0.63) | (0.34) | (0.23) | (0.32) | (0.16) | (0.13) | (0.19) | (0.09) |
| Geography | Citywide | Outer Boroughs | Manhattan | Citywide | Outer Boroughs | Manhattan | Citywide | Outer Boroughs | Manhattan |
| Adjusted $R^2$ | 0.10 | 0.10 | 0.17 | 0.08 | 0.09 | 0.17 | 0.05 | 0.05 | 0.17 |
| *N* | 1,414 | 1,258 | 156 | 1,414 | 1,258 | 156 | 1,414 | 1,258 | 156 |

**B. Deaths - tracts between .2 and 2km from highway**

| | (1) | (2) | (3) | (4) | (5) | (6) | (7) | (8) | (9) |
|---|---|---|---|---|---|---|---|---|---|
| | $PM_{2.5}$ | $PM_{2.5}$ | $PM_{2.5}$ | $NO_2$ | $NO_2$ | $NO_2$ | NO | NO | NO |
| AQ | 0.90** | 1.09** | −0.29 | 0.47** | 0.58** | −0.11 | 0.27* | 0.29* | −0.06 |
| | (0.45) | (0.53) | (0.31) | (0.23) | (0.28) | (0.14) | (0.14) | (0.17) | (0.08) |
| Geography | Citywide | Outer Boroughs | Manhattan | Citywide | Outer Boroughs | Manhattan | Citywide | Outer Boroughs | Manhattan |
| Adjusted $R^2$ | 0.16 | 0.17 | 0.20 | 0.13 | 0.15 | 0.19 | 0.11 | 0.15 | 0.19 |
| *N* | 1,601 | 1,458 | 143 | 1,601 | 1,458 | 143 | 1,601 | 1,458 | 143 |

**C. Deaths - pollution measured at closest monitor**

| | (1) | (2) | (3) | (4) | (5) | (6) | (7) | (8) | (9) |
|---|---|---|---|---|---|---|---|---|---|
| | $PM_{2.5}$ | $PM_{2.5}$ | $PM_{2.5}$ | $NO_2$ | $NO_2$ | $NO_2$ | NO | NO | NO |
| AQ | 0.65* | 1.28** | −0.14 | 0.27* | 0.49** | −0.07 | 0.19* | 0.38* | −0.04 |
| | (0.34) | (0.64) | (0.17) | (0.14) | (0.24) | (0.08) | (0.10) | (0.21) | (0.04) |
| Geography | Citywide | Outer Boroughs | Manhattan | Citywide | Outer Boroughs | Manhattan | Citywide | Outer Boroughs | Manhattan |
| Adjusted $R^2$ | 0.10 | 0.001 | 0.20 | 0.12 | 0.07 | 0.20 | 0.04 | −0.16 | 0.20 |
| *N* | 1,407 | 1,254 | 153 | 1,407 | 1,254 | 153 | 1,407 | 1,254 | 153 |

**D. Deaths - top and bottom 5% windsorized by tract population**

| | (1) | (2) | (3) | (4) | (5) | (6) | (7) | (8) | (9) |
|---|---|---|---|---|---|---|---|---|---|
| | $PM_{2.5}$ | $PM_{2.5}$ | $PM_{2.5}$ | $NO_2$ | $NO_2$ | $NO_2$ | NO | NO | NO |
| AQ | 0.68 | 0.97* | −0.12 | 0.33 | 0.47* | −0.06 | 0.21* | 0.30 | −0.04 |
| | (0.42) | (0.53) | (0.34) | (0.20) | (0.27) | (0.15) | (0.13) | (0.18) | (0.08) |
| Geography | Citywide | Outer Boroughs | Manhattan | Citywide | Outer Boroughs | Manhattan | Citywide | Outer Boroughs | Manhattan |
| Adjusted $R^2$ | 0.19 | 0.17 | 0.20 | 0.18 | 0.17 | 0.20 | 0.16 | 0.13 | 0.20 |
| *N* | 1,309 | 1,167 | 141 | 1,309 | 1,167 | 141 | 1,309 | 1,167 | 141 |

Table S14: Additional Robustness Checks (continued)

| | **E. Deaths - Wind speed excluded as instrument** | | | | | | | | |
|---|---|---|---|---|---|---|---|---|---|
| | (1) | (2) | (3) | (4) | (5) | (6) | (7) | (8) | (9) |
| | $PM_{2.5}$ | $PM_{2.5}$ | $PM_{2.5}$ | $NO_2$ | $NO_2$ | $NO_2$ | NO | NO | NO |
| AQ | 1.02 | 0.97 | 11.96 | 0.39 | 0.39 | −0.32 | 0.20 | 0.21 | −0.16 |
| | (0.89) | (0.82) | (84.48) | (0.37) | (0.39) | (0.85) | (0.19) | (0.21) | (0.41) |
| Geography | Citywide | Outer Boroughs | Manhattan | Citywide | Outer Boroughs | Manhattan | Citywide | Outer Boroughs | Manhattan |
| Adjusted $R^2$ | 0.11 | 0.17 | −46.01 | 0.13 | 0.18 | 0.18 | 0.13 | 0.17 | 0.19 |
| $N$ | 1,407 | 1,254 | 153 | 1,407 | 1,254 | 153 | 1,407 | 1,254 | 153 |
